# Strategies for optimising early detection and first response management of postpartum haemorrhage at caesarean birth: A modified Delphi-based international expert consensus

**DOI:** 10.1101/2023.09.08.23295283

**Authors:** V Pingray, CR Williams, F Alwy Al-beity, E Abalos, S Arulkumaran, A Nabhan, A Blumenfeld, B Carvalho, C Deneux-Tharaux, S Downe, A Dumont, MF Escobar Vidarte, C Evans, S Fawcus, H Galadanci, GJ Hofmeyr, C Homer, A Lewis, T Liabsuetrakul, P Lumbiganon, E Main, F Muriithi, I Nunes, J Ong’ayi, V Ortega, T Phan, Z Qureshi, C Sosa, H Tuyet, J Varallo, A Weeks, M Widmer, OT Oladapo, I Gallos, A Coomarasamy, S Miller, F. Althabe

**Affiliations:** Institute for Clinical Effectiveness and Health Policy, Department of Mother and Child Health Research; Department of Obstetrics & Gynaecology Muhimbili University of Health and Allied Science, Dar es salaam, Tanzania; Maternidad Martin, Secretaria de Salud Publica de Rosario; St George’s, University of London; Institute for Clinical Effectiveness and Health Policy; Stanford University Medical Center; Institut national de la sante et de la recherche medicale (Inserm); UCLan, UK; Universite Paris Cite, IRD, Inserm, Ceped, F-75006 Paris, France; Departamento de Ginecologia y Obstetricia, Fundacion Valle del Lili, Cali 760032, Colombia; Jhpiego, Technical Leadership & Innovations Office; Department of Obstetrics and Gynaecology, University Cape Town; Department of Obstetrics and Gynaecology, Aminu Kano Teaching Hospital; Hung Vuong Hospital, Obstetrics and Gynecology, Ho Chi Minh, VN; Department of Obstetrics and Gynaecology, University of Botswana, Botswana; Burnet Institute; Amherst College; Department of Epidemiology, Faculty of Medicine, Prince of Songkla University, Hat Yai, Songkhla, Thailand; Department of Obstetrics and Gynaecology, Faculty of Medicine, University Khon Kaen; Department of Obstetrics and Gynecology, Maternal Fetal Medicine, Standford University; Liverpool School, Kenya; Institute of Metabolism and Systems Research, University of Birmingham; Department of Obstetrics and Gynecology, Faculty of Medicine, Ain Shams University; Centro Hospitalar Vila Nova de Gaia Espinho, Department of Obstetrics and Gynaecology, Porto, Portugal; Department of Delivery, Tu Du Hospital, Ho Chi Minh City, Vietnam; University of Nairobi; Woman and Reproduction Health Unit at Maternal Health at the Latin American Center of Perinatology (CLAP/WR), PAHO; Global Surgery Foundation, Women’s Health; Department of Women’s and Children’s Health, University of Liverpool. UK; UNDP-UNFPA-UNICEF-WHO-World Bank Special Programme of Research, Development and Research Training in Human Reproduction (HRP), Department of Sexual and Reproductive Health and Research, World Health Organization, Geneva, Switzerland; University of California San Francisco

**Author notes:** **Corresponding author: Veronica Pingray**, Institute for Clinical Effectiveness and Health Policy. These authors contributed equally (V Pingray, CR Williams). SM and FA share senior co-authorship.

**Keywords:** Postpartum haemorrhage, caesarean birth, early detection, first response management, expert consensus

## Abstract

**OBJECTIVE:** There are no globally agreed upon strategies on early detection and first response management of postpartum haemorrhage during and after caesarean birth. Our study aimed to develop an international expert’s consensus on evidence-based approaches for early detection and first response management of PPH intraoperatively and postoperatively in caesarean birth.

**DESIGN:** Systematic review and three-stage modified-Delphi expert consensus.

**SETTING:** International.

**POPULATION:** Panel of 22 global experts in postpartum haemorraghe with diverse backgrounds, and gender, professional, and geographic balance.

**OUTCOME MEASURES:** Agreement or disagreement on strategies for early detection and first response management of postpartum haemorrhage at caesarean birth.

**RESULTS:** Experts agreed that the same PPH definition should apply to both vaginal and caesarean birth. For the intraoperative phase, the experts agreed that early detection should be accomplished via quantitative blood loss measurement, complemented by monitoring the woman’s haemodynamic status; and that first response should be triggered once the woman loses at least 500 mL of blood with continued bleeding or when she exhibits clinical signs of haemodynamic instability, whichever occurs first. For the first response, experts agreed on immediate administration of uterotonics and tranexamic acid, examination to determine aetiology, and rapid initiation of cause-specific responses. In the postoperative phase, the experts agreed that caesarean birth-related PPH should be detected primarily via frequently monitoring the woman’s haemodynamic status and clinical signs and symptoms of internal bleeding, supplemented by cumulative blood loss assessment performed quantitatively or by visual estimation. Postoperative first response was determined to require an individualised approach.

**CONCLUSION:** These agreed-upon proposed approaches could help improve the detection of PPH in the intra and postoperative phases of caesarean birth and the first response management of intraoperative PPH. Determining how best to implement these strategies is a critical next step.

**STRENGTHS AND LIMITATIONS OF THIS STUDY:** *Strengths:*

- Use of a rigorous and systematic process to identify and synthesise high-quality PPH evidence in the literature.
- The selection of the expert panellists ensured a wide range of perspectives to enhance the utility and applicability of this consensus to a wide range of clinical settings.
- There was a very low rate of loss to follow-up and the first two rounds of the modified Delphi process were blinded to avoid social acceptability bias, and the hybrid meeting was facilitated to ensure that all panellists had equal opportunity to contribute to the discussion.

*Limitations:*

- Due to the dearth of quality evidence on PPH related to caesarean birth, experts often had to extrapolate from evidence on interventions recommended for PPH in vaginal birth or make decisions based on their experiences. This sometimes led to omitting interventions that might be useful for early detection or first-response management.
- Given the highly technical content, we did not include recipients of these interventions, or their representatives, among the panellists.
- Since our systematic review, three updated PPH guidelines have been published, with some guidance relevant to PPH during or after caesarean birth. They mostly align with previously published guidance included in our study, with the exception of an increased focus on concealed haemorrhage assessment and one guideline recommending the use of prophylactic tranexamic acid for women at high PPH risk.

## INTRODUCTION

Deaths from postpartum haemorrhage (PPH), the leading direct cause of maternal mortality globally, are potentially preventable with timely diagnosis and management (1,2). The risk of PPH is significantly higher with caesarean birth than vaginal birth, especially in cases of emergency caesarean birth (3). With global caesarean birth rates increasing, PPH during and after caesarean birth is a growing concern (4). The impact is particularly acute in low and middle-income countries (LMICs), where 32% of all maternal deaths after caesarean birth are related to PPH (5). In some LMICs, caesarean births outnumber vaginal births (6). Several factors challenge effective response to PPH in LMICs. These countries have well-documented difficulties accessing surgical services, skilled staff, and blood/blood products (7). Even when access concerns are addressed, use of interventions to detect and manage PPH are often inconsistent (8,9).

A standardised approach to PPH management has been shown to improve outcomes, including significantly reducing severe PPH rates amongst women giving birth vaginally (10). Similarly, studies including women having caesarean birth suggest a reduction in severe morbidity associated with the use of comprehensive haemorrhage protocols (11,12). The World Health Organization (WHO) has published and updated recommendations for the prevention and treatment of PPH (2,13,14). However, these recommendations neither detail methods for early detection of PPH during and after caesarean birth nor clearly indicate when to initiate treatment (i.e., the ‘trigger’ criteria), both of which may contribute to observed variations in clinical practice (2,7,15). PPH management practices may vary depending on whether the haemorrhage occurs during or after the surgical procedure (16). Proposing standardised and evidence-based global strategies may help to reduce practice variations and improve the quality of care. Our study aimed to develop an international consensus on standardised approaches for PPH detection and first response management during and after caesarean birth.

## METHODS

The study involved a systematic review and an expert consensus using a three-stage modified-Delphi process.

### Systematic review

A systematic review of published national and international guidelines for PPH prevention and management was conducted to identify interventions for collecting and measuring blood loss, methods for detecting PPH, thresholds for treatment, and first response interventions to manage PPH both during surgery (intraoperative) and after surgery (postoperative). To be included, the guidelines needed to include guidance on the detection or management of PPH during or after caesarean birth. The literature search in PubMed, EMBASE, CINAHL, and Cochrane Library databases included papers published from January 2012 to July 2022 (Supplementary File S1). The search was complemented by reviewing the English-language grey literature to identify guidelines.

Since few of these guidelines were focused specifically on the intra- or postoperative phases or described PPH detection methods, an additional systematic search was conducted, focused on PPH detection and management during and after caesarean birth. Peer-reviewed systematic reviews of RCTs were eligible. Subject matter experts were consulted to add any relevant peer-reviewed articles missed by the systematic search.

Titles and abstracts of both guidelines and systematic reviews of RCTs were screened by pairs of independent reviewers who subsequently reviewed full texts, conducted quality appraisals, and extracted data using previously piloted forms. Only guidelines with AGREE II scores between 5 and 7 and systematic reviews with modified-AMSTAR quality assessment of “Moderate” or “High” were eligible for data extraction (17,18). The results of the systematic review were used to inform the development of the Delphi surveys and to provide the experts with summaries of the existing evidence.

### Expert consensus

A three-stage modified Delphi process was conducted between December 2021 and September 2022, with two rounds of individual online surveys, followed by a third round: a hybrid (virtual and in-person) meeting with group discussions and final voting. Twenty-five PPH experts with the knowledge and skills to critically assess scientific evidence were invited to participate in all three rounds. They included specialists in nursing, midwifery, obstetrics, surgery, and anaesthesia. The experts were selected to ensure gender, professional, and geographic balance. Most experts were co-authors of recent national and international guidelines or principal or co-investigators of PPH clinical trials. The same experts were invited to participate in all three rounds. In the third round, observers representing professional associations and WHO regional offices, or who were leaders in PPH research were invited to share their views, but were not eligible to vote.

Based on the findings of the systematic review, questionnaires with open- and close-ended questions were developed, piloted, and administered using Survey Monkey™. A summary of the themes and interventions included in the surveys and criteria used to guide judgements are described in Box 1. The criteria, methods, interventions, and other items included in the surveys were presented with definitions to facilitate interpretation. The themes were explored separately for the intraoperative and postoperative phases. Experts were asked to consider the postoperative PPH phase as only the first two hours immediately after the operation. Each online survey was available for response for six weeks, and three reminders were sent to participants with incomplete or no responses. In the first round, experts were asked to rate caesarean-related PPH definitions, detection methods, thresholds to trigger treatments, and first response interventions. In the second round, experts received their previous individual ratings and group rating distributions. They were asked to re-rate detection methods with disagreement, rank-order the thresholds and first response treatments that had previously received high ratings, and rate new questions that emerged from experts’ comments in open-ended questions from Round 1. In the third round, experts met for a two-day hybrid meeting to discuss areas of divergence between surveys’ findings and to rate (anonymously) final sets of interventions. The agenda and questions guide used to facilitate the discussion are available in the Supplementary Materials (Supplementary File S2 and S3). Figure 1 outlines the process of consensus building.

#### Box 1: Themes explored, and criteria used to guide assessments

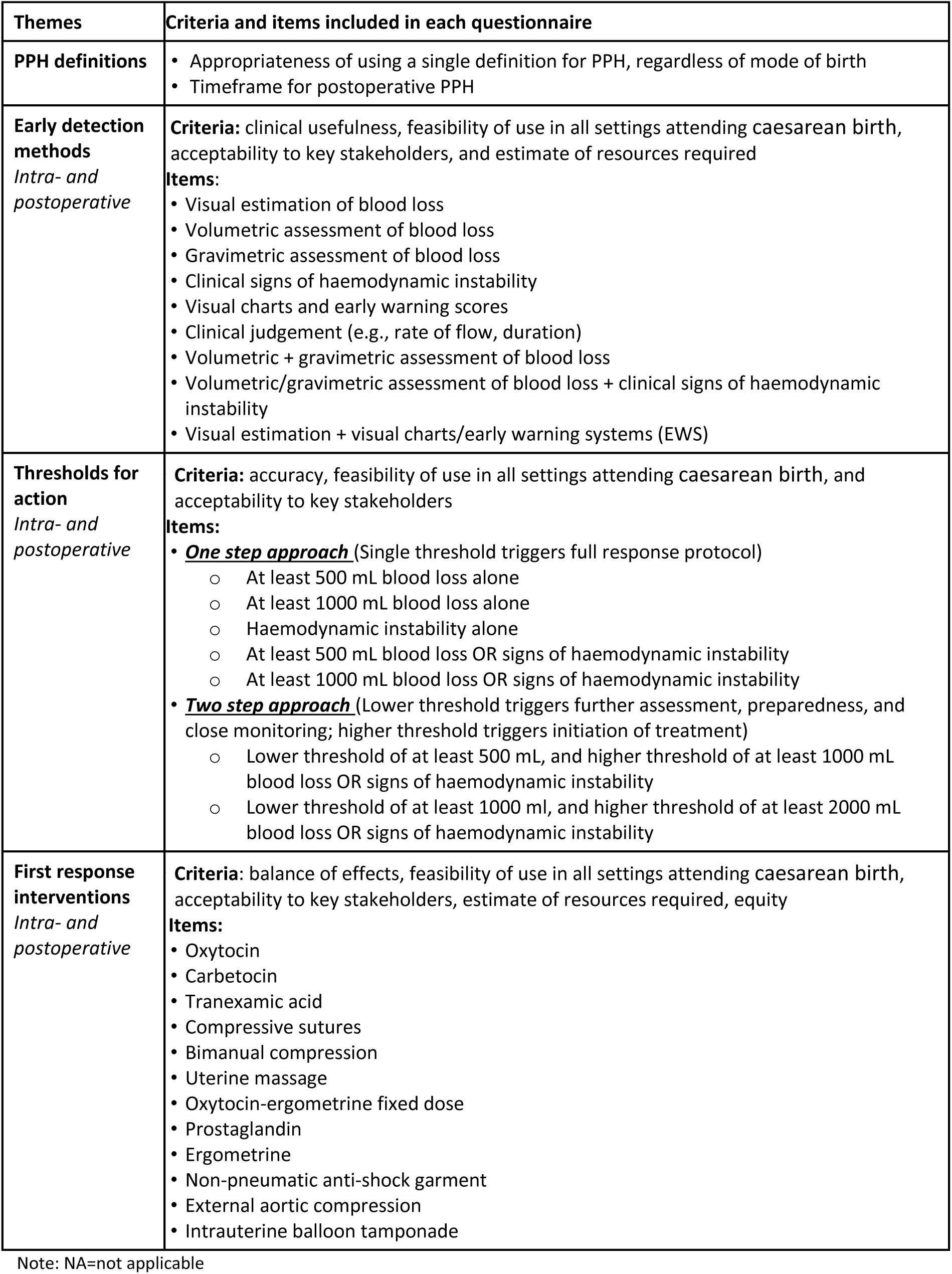

**Figure 1.**
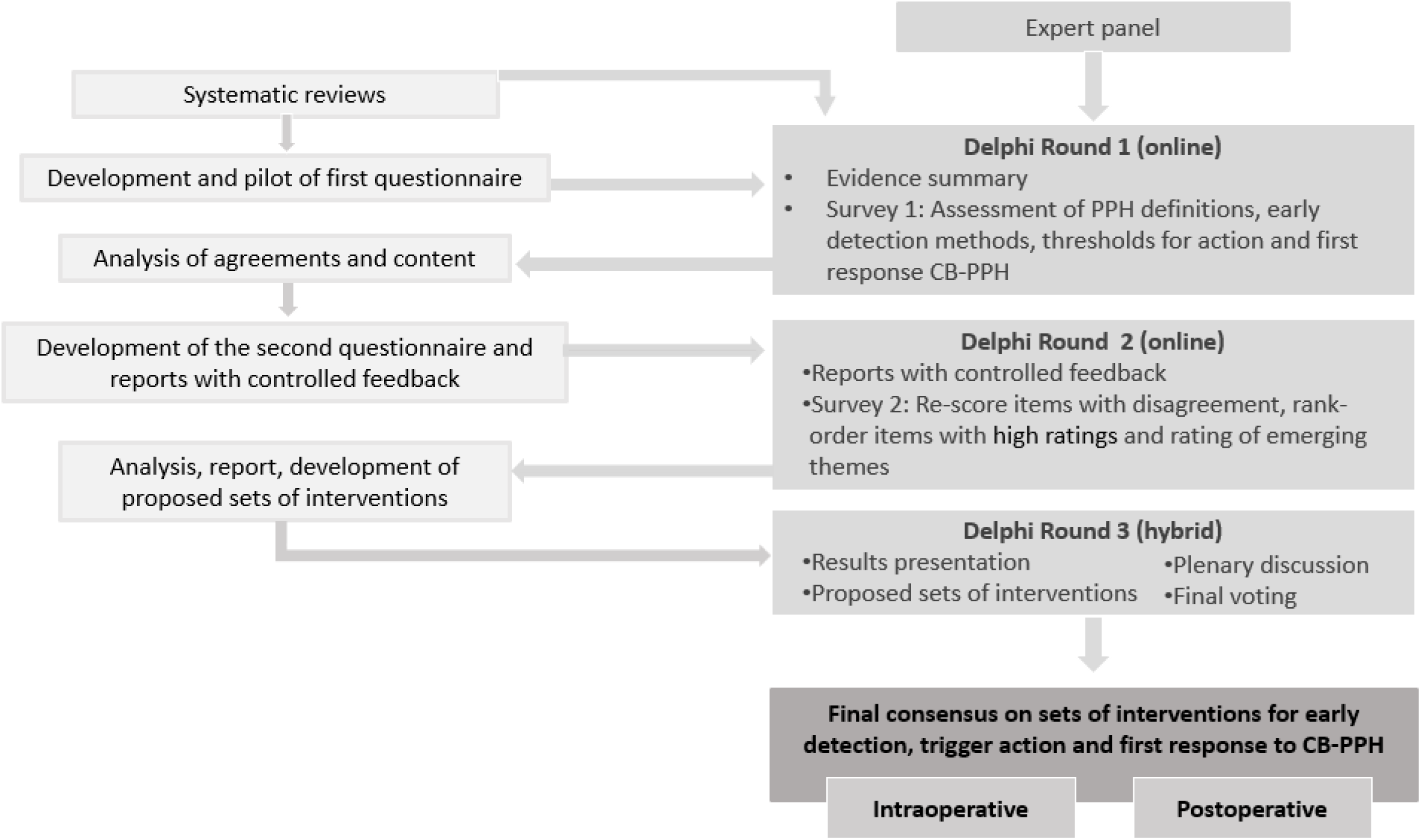
Technical consultation flowchart.

Median group rating and disagreement index (DI) were calculated to summarise experts’ ratings and to measure agreement. A DI < 1 indicated agreement, while a DI ≥ 1 indicated disagreement (19). The RAND/UCLA appropriateness scale was used to classify interventions as “appropriate”, “inappropriate”, or “uncertain” (19). Interventions with median ratings in the top third of the appropriateness scale (7–9) were classified as “appropriate”; those in the bottom third were classified as “inappropriate” (1–3); and those with intermediate median ratings were classified as “uncertain” (4–6). Domains with disagreement among the experts were also classified as “uncertain” (Supplementary Figure S1).

## RESULTS

The systematic search identified 802 guidelines and systematic reviews. After screening and quality appraisal, 17 guidelines (2,13,15,20–33), four systematic reviews (34–37) and 15 peer-reviewed studies were included (38–53) (Supplementary Figure S2). Included guidelines and systematic reviews identified six PPH definitions, five PPH detection methods, ten blood loss collection devices, seven thresholds to initiate treatment, and 14 interventions to conservatively manage PPH. Results are in Supplementary Tables S1-S4.

Of 25 experts invited, 22 agreed to participate in the Delphi process (Supplementary File S4). All completed the first and second rounds, while 20/22 participated and voted in the third round. The experts who completed all rounds were from 11 countries from all WHO world regions (six from the African Region, one from the Eastern Mediterranean Region, three from the European Region, six from the Region of the Americas, two from the South-East Asian Region, and two from the Western Pacific Region). They had different professional backgrounds (obstetricians and gynaecologists, anaesthetists, surgeons, nurse-midwives, and midwives) and were gender-balanced (12 men and 10 women). In addition, four observers participated in the discussion during the third round but did not vote.

The median ratings and measures of agreement obtained from the first and second rounds of online surveys are given in Supplementary Tables S5-S8 and Figure S3. Experts’ ratings and agreements in the third round are given in Table 1. Consensus was reached for (a) using a single definition for PPH, regardless of mode of birth, (b) early detection of PPH at caesarean birth and thresholds to initiate treatment in the intraoperative phase, (c) clinical interventions for first response management of intraoperative PPH, and (d) early detection of PPH after caesarean birth and thresholds to initiate treatment in the postoperative phase. However, the first response treatment in the postoperative phase was determined to require an individualised approach.

**Table 1.** Experts’ ratings and agreement on early detection and first response to intraoperative and postoperative PPH.

| Set of clinical interventions | Rating distribution |  |  | Agreement |  |  |  |
| --- | --- | --- | --- | --- | --- | --- | --- |
|  | # votes in each interval |  |  | Median (IQR) in a<br>1 - 9 differential<br>scale | RAND DI | Qualitative<br>scale | Appropriateness<br>scale |
|  | 1 - 3 | 4 - 6 | 7 - 9 |  |  |  |  |
| **Intraoperative** |  |  |  |  |  |  |  |
| *Early detection of PPH in all women having caesarean birth* |  |  |  |  |  |  |  |
| Quantitative blood loss measurement and monitoring of haemodynamic status | 0 | 1 | 19 | **8 (1)** | **-0.34** | **Yes** | **Appropriate** |
| *Thresholds for triggering action* |  |  |  |  |  |  |  |
| Option 1: At least 500 mL with continued bleeding OR clinical signs of haemodynamic instability | 1 | 3 | 16 | **8 (1)** | **-0.13** | **Yes** | **Appropriate** |
| Option 2: At least 750 mL with continued bleeding OR clinical signs of haemodynamic instability | 7 | 4 | 9 | 6 (5) | 3.13 | No | Uncertain |
| **Postoperative** |  |  |  |  |  |  |  |
| *Early detection of PPH in all women having caesarean birth* |  |  |  |  |  |  |  |
| Monitoring of haemodynamic status and quantitative blood loss measurement if feasible | 0 | 2 | 18 | **8 (1)** | **-0.34** | **Yes** | **Appropriate** |
| *Thresholds for triggering action* |  |  |  | Not applicable |  |  |  |

Notes: <sup>a</sup>RAND DI: The disagreement index is a continuous scale used to measure the dispersion of experts' ratings, taken as an indicator of the level of agreement; <sup>b</sup>Agreement: A DI < 1 represents an agreement, while a DI ≥ 1 indicates disagreement; <sup>c</sup>Appropriateness: Items are classified as "appropriate" with median ratings in the top (median between 7-9) third and agreement, and as "uncertain" with intermediate median ratings (median between 4-6) or with disagreement.

### Definition of PPH during and after caesarean birth

The experts agreed that a single definition of PPH, regardless of mode of birth (median rating 7.5; DI −5.23). Specifically, they agreed that the definition of PPH during and after caesarean birth should be the same as the definition of PPH related to vaginal birth, to underscore the importance of rapid action to address excessive bleeding.

### Intraoperative phase

#### Early detection of PPH during caesarean birth and thresholds for triggering action

Experts agreed that during caesarean birth, blood loss should be assessed via quantitative measurement, complemented by ongoing monitoring of the woman’s haemodynamic status (median rating 8; DI −0.34). Further, quantitative measurement and monitoring should be conducted for all women having a caesarean birth (Box 2). They noted the importance of distinguishing blood from amniotic fluid. This might be achieved by using separate suction canisters or measuring and recording the amount of amniotic fluid within the canister immediately after the birth and before delivery of the placenta.

##### Box 2: Agreed early detection of PPH during caesarean birth and thresholds for triggering first response in the intraoperative phase

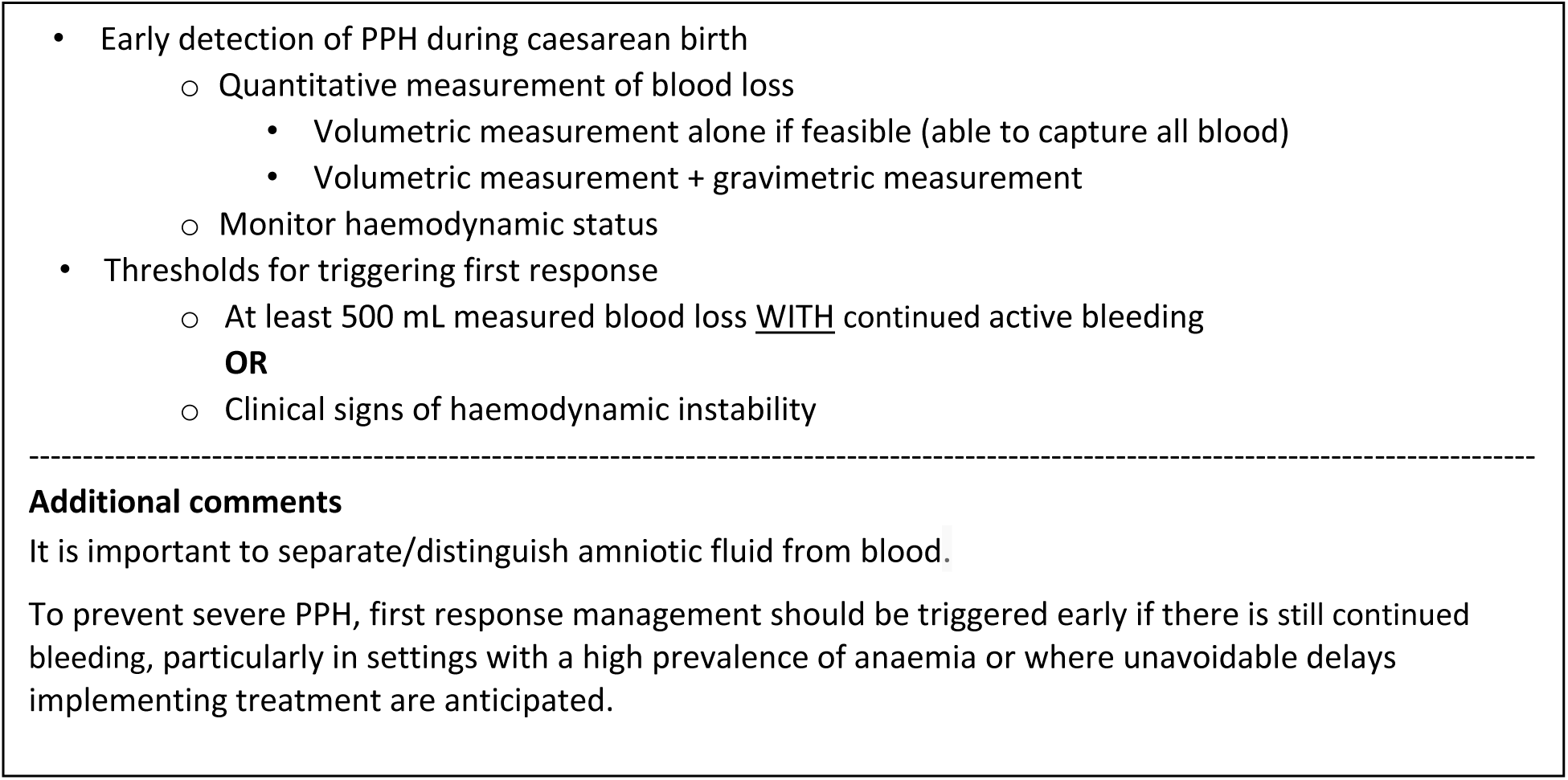

The experts agreed that first response treatment should be triggered if the woman has lost at least 500 mL blood and still has continued bleeding or if she exhibits clinical signs of haemodynamic instability, whichever occurs first (median rating 8; DI −0.13). Such early action was considered important to prevent severe PPH and associated morbidity, because measurement of blood loss lags actual blood loss. Rapid response has been identified as a critical component of the effectiveness of an early detection and PPH treatment strategy to prevent severe PPH in vaginal births (40). Experts considered that rapid response is particularly important in settings with high prevalence of anaemia. It was noted that the proposed threshold for triggering action may result in many women receiving first response treatment for PPH. Some experts pointed out that this could diminish providers’ responsiveness and recognition of PPH as a serious complication.

#### First response management: intraoperative phase

The agreed first response management is summarised in Box 3. Specifically, the experts agreed clinicians should commence an infusion of oxytocin. If a prophylactic or other oxytocin infusion is already in place, the anaesthetist should quickly maximise the oxytocin dose as increasing uterine tone helps to reduce bleeding from the incision. If atony is diagnosed or the bleeding continues, the anaesthetist should rapidly add in a different uterotonic for treatment. The experts noted that this should occur quickly, rather than waiting to see whether the bleeding is responsive to oxytocin. They also agreed that tranexamic acid (TXA) should be administered as first response treatment, unless the woman had already received TXA within the last 30 minutes. Next, the team should carefully examine the woman to determine the source(s) of bleeding and initiate a cause-specific response. If the bleeding is due to trauma, the surgical team should close the uterus, repair any tears, and attend to the wound. If the bleeding is due to uterine atony, the surgical team should control bleeding mechanically with intra-abdominal uterine massage or massage the exteriorised uterus, as the anaesthetic team manages uterotonic administration, as previously described. Some experts noted that the assessment of atonic PPH may require the lifting of surgical drapes to assess vaginal blood loss. Experts highlighted that bleeding may be due to a combination of trauma and uterine atony; in such cases the team should take a comprehensive approach. The experts also highlighted the importance of exteriorising and examining the posterior side of the uterus for tears and occult uterine rupture.

##### Box 3: Agreed upon first response treatment for PPH during the intraoperative phase

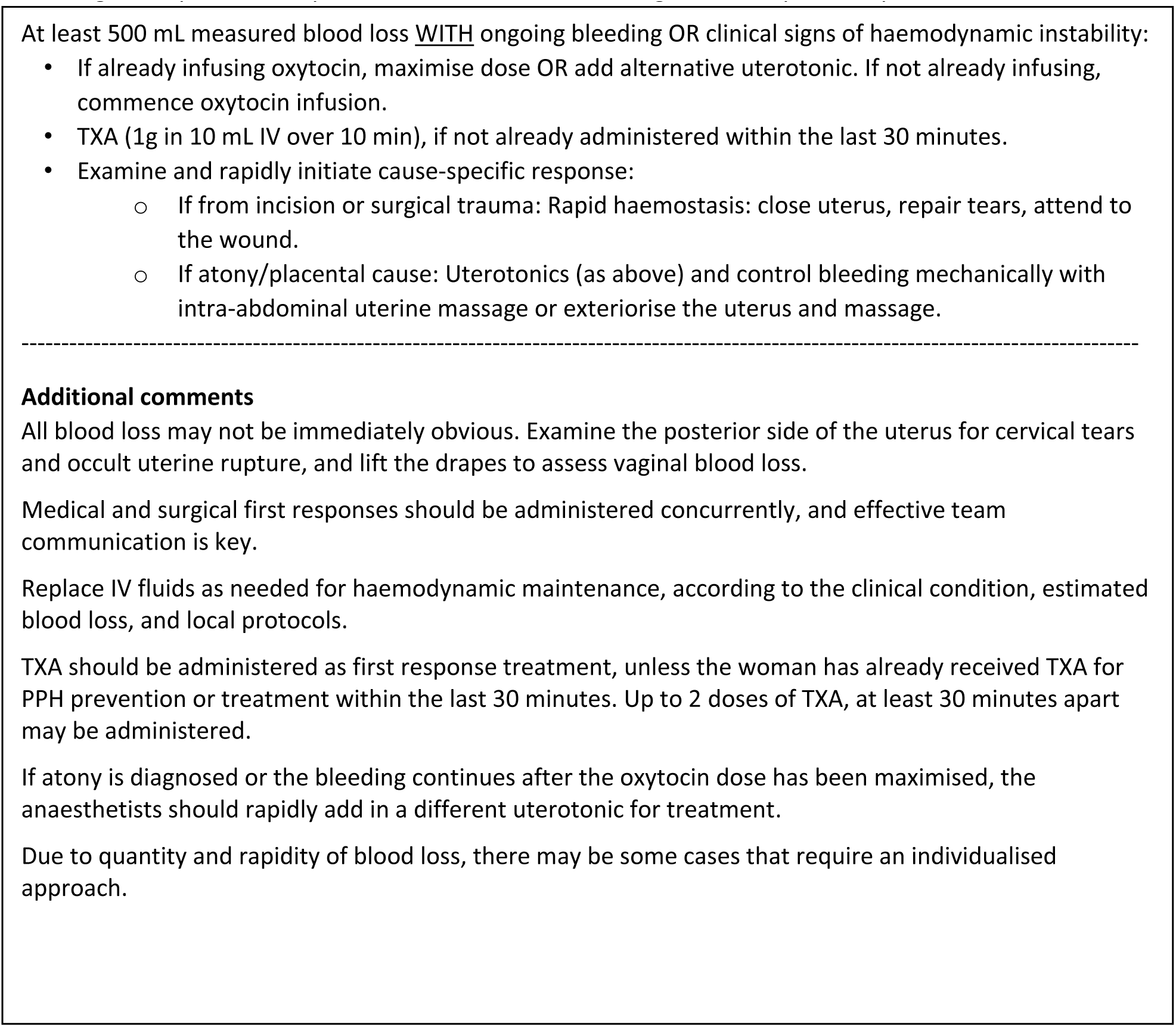

Surgical and anaesthetic teams should mobilise to administer the surgical and medical first responses concurrently. Team communication can be challenging and should be practised in drills to develop effective messages that will not alarm women. Teams should immediately call for senior assistance when necessary.

Experts also noted that anaesthetic teams should give IV fluids as needed for haemodynamic maintenance, according to the clinical condition and estimated blood loss. The literature is unclear regarding which specific fluids should be used, the optimal volumes, and how best to monitor haemodynamic status. Until further evidence is available, experts advised each setting to develop or follow local protocols.

Finally, experts acknowledged that this first response approach is intended to be appropriate for most cases of intraoperative PPH. There may be some cases that, due to quantity and rapidity of blood loss, require an individualised approach.

### Postoperative phase

#### Early detection of PPH after caesarean birth and thresholds for triggering action

Postoperative detection of PPH based on monitoring blood loss can be misleading because of internal bleeding. Thus, during this phase, experts agreed (median rating 8, DI −0.34) that blood loss should be assessed primarily through frequent monitoring of women’s haemodynamic status (when possible, at least every 15 minutes for the first 2 hours) and clinical signs and symptoms of internal bleeding (e.g., assessment of fundal height) (Box 4). Some experts noted that postoperative monitoring for at least 30 minutes after caesarean birth should occur in a designated recovery area to ensure the woman’s safety. If internal bleeding is suspected, experts recommended an urgent ultrasound assessment if available. In addition, if the assessment of postoperative vaginal blood loss is feasible, either by quantitative measurement or estimation (e.g., counting pads), it should be performed. Experts agreed that, when possible, measured postoperative blood loss should be added to the quantified intraoperative blood loss, though they acknowledged that this may be challenging in some settings.

##### Box 4: Agreed early detection of postoperative PPH and thresholds for triggering first response

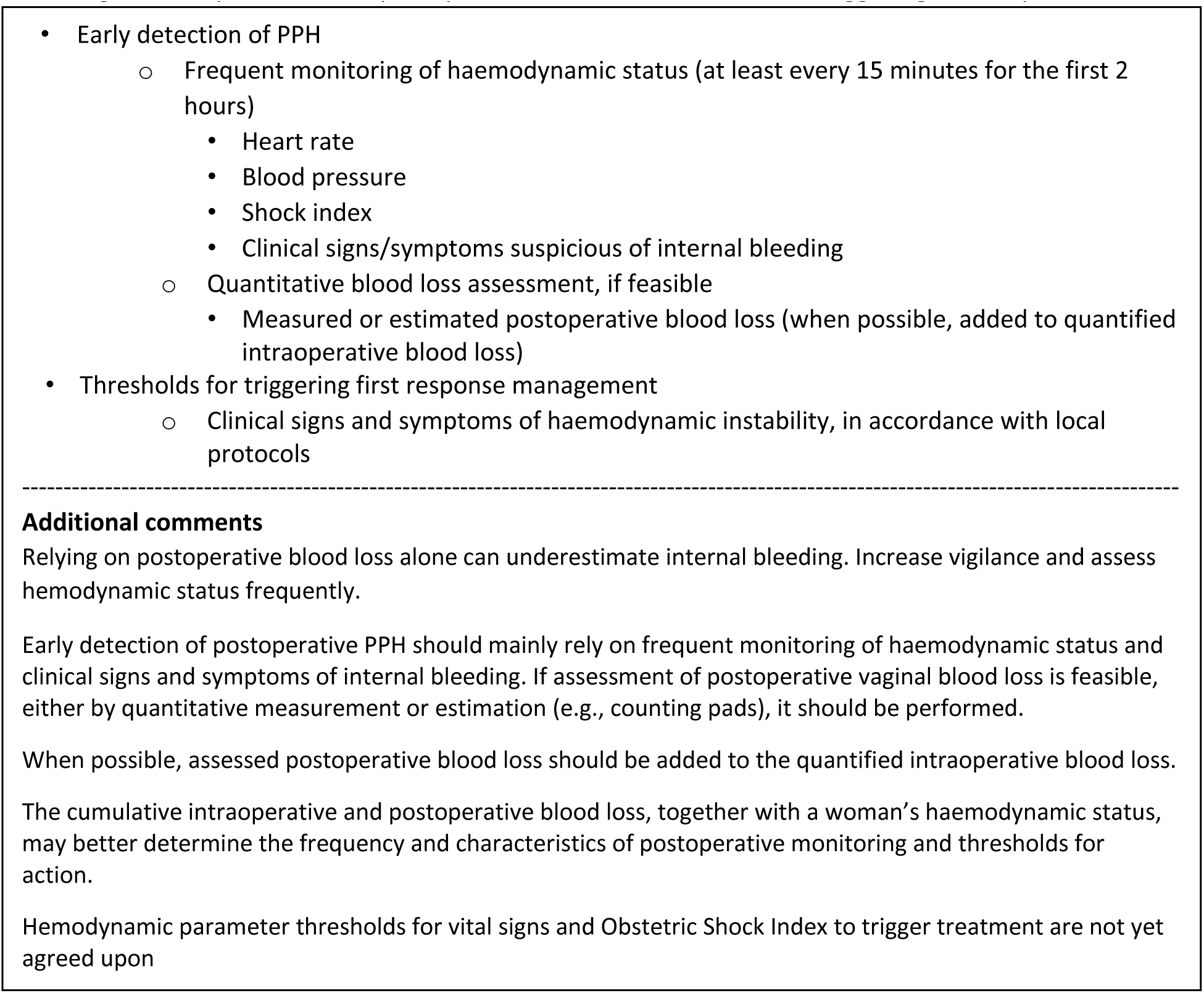

Experts noted that cumulative intraoperative and postoperative blood loss, together with a woman’s haemodynamic status, can help adjust the frequency and characteristics of postoperative monitoring and thresholds for action. For example, a woman who experienced substantial blood loss intraoperatively may require more frequent monitoring than the baseline every 15 minutes.

The experts acknowledged that haemodynamic parameters for postoperative thresholds to trigger treatment are still unclear. Until further evidence is available, each setting should develop or follow their local protocols. The Obstetric Shock Index (heart rate divided by systolic blood pressure; OSI) has been used in some settings, but there is not yet clear evidence on appropriate cut-off points.

#### Postoperative phase: First response management

The experts noted that the follow-on postoperative treatment approach may vary substantially according to many factors, including the woman’s baseline risk, anaemia, whether intraoperative PPH occurred, the woman’s postoperative haemodynamic status, and clinical signs and symptoms of internal bleeding (e.g., assessment of fundal height; if available, ultrasound, paracentesis). Until further evidence is available, experts recommended that local protocols be developed that consider these factors, rather than relying on a common postoperative first response approach for all cases and settings.

#### Experts’ final comments

The experts recognised that detection methods and first response interventions for PPH are essential for the care of all women having a caesarean birth, regardless of their risk status. However, women at high risk of developing PPH may require additional specialised monitoring and care.

In addition, given that PPH can arise intra- or postoperatively for any woman, strategies for early detection of PPH should be incorporated into routine practice alongside PPH prevention and risk assessment.

Finally, experts highlighted two cross-cutting remarks regarding PPH during and after caesarean birth. First, good surgical practices, as recommended by the WHO Guidelines for Safe Surgery, should be followed to prepare for, perform, and follow-up caesarean births (54). The routine use of WHO surgical safety checklists have proven beneficial in reducing perioperative complications (55). Second, it was noted that teamwork, communication, and cooperation are critical. Effectively implementing the early detection and first response interventions described will require training, supportive supervision, monitoring, and evaluation.

## DISCUSSION

### Main findings

Expert consensus on optimal approaches for detecting and managing PPH during and after caesarean birth was developed among an international panel. Through two systematic reviews and a three-round modified Delphi process, consensus was reached for (a) using a single definition for PPH, regardless of the mode of birth, (b) early detection of PPH during caesarean birth and thresholds to initiate treatment in the intraoperative phase, (c) clinical interventions for first response to intraoperative PPH, and (d) early detection of PPH after caesarean birth and threshold to initiate treatment in the postoperative phase. First response treatment in the postoperative phase was determined to require an individualised approach.

### Strengths and limitations

Study strengths include the use of a rigorous and systematic process to identify and synthesise PPH evidence in the literature. We conducted in-depth systematic reviews with detailed quality appraisals to ensure that we used only high-quality evidence to identify approaches for PPH detection and management interventions. The selection of the expert panellists ensured a wide range of perspectives, to enhance the utility and applicability of this consensus to a wide range of clinical settings. There was a low rate of loss to follow-up. The first two rounds of the modified-Delphi process were blinded, to avoid social acceptability bias, and the hybrid meeting was facilitated by members of the Steering Group, to ensure that all panellists had equal opportunity to contribute to the discussion. The staged modified-Delphi process allowed ample time for discussion and input, and experts provided additional comments to refine the final statements for clarity and accuracy.

Limitations included a dearth of quality evidence on PPH related to caesarean birth. Despite ample evidence on PPH during and after vaginal birth, there is far less published evidence on caesarean birth. Often, the experts had to extrapolate from evidence on interventions recommended for PPH in vaginal birth and make decisions based on their experiences, expert opinions, and best practices, rather than evidence from comparative research. In some cases, this led to omitting interventions that might be useful for early detection or first-response management because there was no rigorous evidence available. It is also a limitation that, given the highly technical content, we did not include recipients of these interventions, or their representatives, among the panellists.

Additionally, since this systematic review of guidelines was conducted, three updated PPH guidelines have been published (56–58). None of these guidelines are specific to PPH at caesarean birth, though all contain some guidance relevant to PPH during or after caesarean birth. The recommendations within these guidelines generally align with previously published guidance included in our study, with a few notable exceptions. The revised 2023 FIGO PPH guideline recommends the use of the Obstetric Shock Index (with a threshold of ≥0.9 triggering first-response treatment), together with the rule of 30, while acknowledging that “the association between shock parameters and advanced treatment modalities in severe PPH has yet to be reported” (59). In the updated CMQCC Obstetric Hemorrhage Toolkit, greater emphasis is placed on assessing for concealed haemorrhage. The guideline recommends using a combination of clinical signs of hypovolemia, the SI, and Early Warning Score to enable earlier postoperative PPH detection (57). The Royal College of Physicians of Ireland guideline suggests that prophylactic tranexamic acid administration be considered in women at high PPH risk (58). The timing of our study prevented us from incorporating these revisions into our systematic review.

### Interpretation

This expert consensus aligns with the recent expert consensus developed by the African Perioperative Research Group (APORG) Caesarean Delivery Haemorrhage Group (60) for clinicians working in Africa. The APORG expert consensus had a broader scope, encompassing antenatal and perioperative prevention, preparedness, first response, and refractory treatment interventions, as well as community- and health system-level indirect interventions. This present expert consensus focuses only on early detection and first response, including specific thresholds for triggering action.

With rates of caesarean birth rising globally, particularly in middle-income countries (6), this research is timely and crucial. International initiatives are underway to end preventable deaths due to PPH, such as the Roadmap to Combat Postpartum Haemorrhage between 2023 and 2030 (61), and the Pan American Health Organization’s Zero Maternal Deaths by Hemorrhage campaign (62). The present expert consensus on early detection and first-response treatment for PPH at caesarean birth adds to existing efforts by clearly delineating how interventions need to be tailored for the context of caesarean birth. This consultation represents an important first step toward developing standardised strategies for reducing morbidity and mortality related to PPH during and after caesarean birth. Determining how best to implement these standardised strategies is a critical next step.

Insights from implementation science suggest that defining evidence-based interventions is a necessary but insufficient step towards changing clinical practice (63). Establishing implementation approaches is believed to increase uptake and fidelity of evidence-based interventions (64). Clinical bundles are one implementation approach that has gained traction in recent years (56,65–68). Global evidence suggests that clinical bundles are a powerful implementation approach for early detection and first response for PPH after vaginal birth (68,69). However, it is unclear whether a bundle is the most appropriate implementation approach for PPH during and after caesarean birth. Bundles require a set of interventions to be administered together, but the administration of some of the clinical interventions outlined here may depend on what occurs during surgery and what other interventions may already have been administered. As such, other implementation approaches, such as algorithms, protocols, or checklists, might be more appropriate (69,70). Defining the optimal implementation approach for early detection and first response management of PPH during and after caesarean birth still remains to be completed. Conducting necessary research to answer this question should be an immediate next step.

In addition, efforts should be pursued to agree on standardised approaches for the management of refractory PPH during and after caesarean birth. These standardised approaches should encompass both the specific interventions used to manage refractory PPH and the implementation strategies to support their uptake and sustainability. Standardised approaches will need to be applicable to a variety of settings, including those with limited access to medical specialists.

## CONCLUSION

This expert consensus proposes strategies for early detection and first response to PPH during and after caesarean birth. Future research should determine how best to implement these strategies and evaluate the effectiveness of the proposed implementation approach. Such research should be conducted soon, so that the approaches and interventions proposed here can rapidly be operationalised and institutionalized to contribute to the global efforts to reduce maternal death and disability.

## CONTRIBUTION TO AUTHORSHIP

SM, FA, AC, IG, and OTO conceived the idea. VP developed the protocol with input from SM and FA. SM, FA, CRW, and VP coordinated the project with input from IG and AC. FM, AL, VO, AB, CRW, and VP conducted the systematic review. OTO, IG, AC, EA, SA, NA, FAA, BC, CDT, SD, AD, MFEV, CE, SF, HG, JH, CH, TL, PL, EM, FM, IN, OJ, QPNP, ZQ, CS, JV, AW, FA, SM, CRW, and VP participated in the consensus and provided expert input at various stages of the project. Data were analysed by CRW and VP with input from SM and FA. SM, FA, CRW, and VP wrote the manuscript with input from all co-authors. This manuscript represents the views of the authors and not the views of the World Health Organization or the UNDP/UNFPA/UNICEF/WHO/World Bank Special Programme of Research, Development and Research Training in Human Reproduction (HRP).

## ETHICAL CONSIDERATIONS

Authors confirm that this exercise was conducted according to relevant guidelines and regulations with respect to the Helsinki Declaration. Experts were advised that participation was voluntary and indicated informed consent. Given the consultative nature of the exercise which involved consenting experts in their work capacity, ethical clearance was not required.

## CONSENT FOR PUBLICATION

All authors consented to publication.

## PATIENT AND PUBLIC PARTNERSHIP

Given the highly technical content, we did not include recipients of these interventions, or their representatives, among the panellists. This limitation is addressed in the discussion.

## Data Availability

Extensive data are presented in the supplementary materials. Given the small sample size, datasets will be available upon reasonable request.

## ACKNOWLEDGMENTS

We would like to acknowledge the contribution of Bill & Melinda Gates Foundation officers for conceiving the research question and supporting the conduct of this study; the contribution of Zhitong Yu in preparing the consensus building process and the administrative support of Chastine Sebolino and Maria Harmitton Oliveto.

This article reflects the views of the named authors, and not the views of their organisations, or the views of UNDP-UNFPA-UNICEF-WHO-World Bank Special Programme of Research, Development and Research Training in Human Reproduction (HRP) or World Health Organization.

