## Supplementary materials for "Strategies for optimising early detection and first response management of postpartum haemorrhage at caesarean birth: A modified Delphi-based international expert consensus"

#### Supplementary Tables

**Table S1.** Definitions of PPH and severe PPH used in clinical guidelines

| Definitions | Number of guidelines using the definition | Specific guidelines using the definition | Specifications regarding mode of birth and timeframe in which bleeding occurs | Additional considerations |
|---|---|---|---|---|
| **DEFINITIONS OF PPH** | | | | |
| *Blood loss at least 500 mL* | 6 | (1, 2, 3, 4, 5, 6) | Explicitly specified that this definition applied regardless of the mode of birth: (3, 6)<br>Indicated that blood loss must occur within 24 h of birth: (1, 2, 6) | Two guidelines targeting high-income countries (HICs) specified that for caesarean birth the threshold could be set at a higher blood loss if clinically tolerated: (3, 4) |
| *Blood loss at least 1000 mL* | 2 | (7, 8) | Definition applied for caesarean only: all<br>No timeframe indicated: all | --- |
| *Blood loss at least 1000 mL OR signs of haemodynamic instability* | 5 | (9, 10, 11, 12, 13) | Definition applied for caesarean only: (9, 10)<br>Indicated that blood loss must occur within 24 h of birth: all | One guideline recommended that cumulative blood loss of 500-999 mL alone should trigger increased supervision and potential interventions as clinically indicated: (13) |
| *Any bleeding that causes haemodynamic instability* | 1 | (14) | Explicitly specified that definition was applicable regardless of mode of birth<br>Indicated that bleeding must occur within 24 h of birth | For clinical purposes, any blood loss that had the potential to produce hemodynamic instability should be considered PPH. The amount of blood loss required to cause hemodynamic instability would depend on pre-existing conditions (e.g., anaemia, dehydration, gestational hypertension with proteinuria) |
| **DEFINITIONS OF SEVERE PPH** | | | | |
| *Blood loss at least 1000 mL* | 4 | (1, 2, 3, 5) | Explicitly specified that this definition applied regardless of the mode of birth: (3)<br>Indicated that bleeding must occur within 24 h of birth: (1, 2) | RCOG defined major PPH as blood loss greater than 1000 mL. Major PPH could be further subdivided into moderate (1001–2000 ml) and severe (more than 2000 ml). |
| *Blood loss at least 1000 mL OR signs of haemodynamic instability* | 2 | (6, 12) | Explicitly specified that this definition applied regardless of the mode of birth: (6)<br>Indicated that bleeding must occur within 24 h of birth: all | --- |

Note: No definition of PPH mentioned: (15, 16, 17). No definition of severe PPH mentioned (4, 8, 11, 13, 16, 17, 18)

**Table S2.** Synthesis of evidence and considerations regarding approaches to detect intraoperative and postoperative caesarean PPH and haemodynamic instability

| Detection methods | Paired with any of the following blood collection devices | Sources discussing this method | Summary of the evidence |
|---|---|---|---|
| **BLOOD LOSS ASSESSMENT METHODS WITH SPECIFIC BLOOD COLLECTION DEVICES** | | | |
| *Visual estimation of blood loss* | • Suction canister<br>• Blood-soaked materials and clots<br>• Calibrated drape<br>• Non-calibrated blood loss collectors placed under the buttocks<br>• Non-calibrated blood loss collectors attached to the abdomen during surgery | 4 Guidelines:<br>(1, 8, 11, 14)<br><br>3 Systematic reviews:<br>(19, 20, 21)<br><br>Additional relevant references:<br>(22) | While four guidelines stated that visual estimation was used in practice, none actually recommended this method. Rather, they described its limitations. Visual estimation was described as subjective, imprecise, and known to underestimate actual blood loss (5, 17). Clinicians were advised to be aware that visual estimation of peripartum blood loss is inaccurate (2). The systematic reviews noted that some trialists used visual estimation to determine amount of blood lost and detect PPH. No further comment was provided. Although widely used for the detection of PPH, visual estimation of blood loss was consistently reported as inaccurate (22). While both underestimation and overestimation occur, the extent of underestimation increased as the volume of blood loss increased (17, 22). |
| *Volumetric* | • Calibrated suction canister<br>• Calibrated drape | 6 Guidelines:<br>(2, 3, 4, 6, 13, 17)<br><br>3 Systematic reviews:<br>(19, 20, 21)<br><br>Additional relevant references:<br>(23, 24, 25, 26, 27) | Guidelines described the use of volumetric methods. Some guidelines proposed using a combination of volumetric and gravimetric methods for assessing blood loss (3, 17). Some guidelines noted that while quantitative methods are more accurate than visual estimation in determining maternal blood loss, their effect on clinical outcomes has not been demonstrated (2, 17). The systematic reviews noted that some trialists used this method to determine amount of blood lost and detect PPH. Some trialists used a combination of gravimetric and volumetric methods. No further comment was provided. Volumetric techniques appeared to be more accurate than the visual estimation of blood loss, irrespective of provider experience, level of training, or specialty (23, 24). Mean measured blood loss was found to be 30% more accurate than estimated blood loss in vaginal births(25). The discrepancy between volumetric methods and visual estimation was found to be higher with increasing blood volume (26). The effectiveness of volumetric methods on clinical outcomes has not been demonstrated (17). A large multicentre multi-country cluster randomized trial comparing calibrated drapes vs. visual estimation failed to show a reduction in severe PPH (27) |
| *Gravimetric* | • Non-calibrated blood loss collectors placed under the buttocks<br>• Non-calibrated blood loss collectors attached to the abdomen during surgery<br>• Blood-soaked materials and clots: either intra- or postoperative | 8 Guidelines:<br>(2, 3, 4, 5, 6, 12, 13, 17)<br><br>2 Systematic reviews:<br>(19, 20)<br><br>Additional relevant references:<br>(28, 29) | Guidelines described the use of gravimetric methods. Some guidelines proposed using a combination of volumetric and gravimetric methods for assessing blood loss (3, 12, 17). Some guidelines noted that while quantitative methods are more accurate than visual estimation in determining maternal blood loss, their effect on clinical outcomes has not been demonstrated (2, 12, 17). Two guidelines stated that weighing of swabs *may* be used, but did not directly suggest their use (2, 4). The systematic reviews noted that some trialists used this method to determine amount of blood lost and detect PPH. Some trialists used a combination of gravimetric and volumetric methods. No further comment was provided. A 2014 randomized controlled trial including nine hundred women presenting for vaginal delivery found that blood loss recorded using a non-calibrated collector followed by gravimetric assessment was lower than blood loss recorded using the calibrated drape for |

| Detection methods | Paired with any of the following blood collection devices | Sources discussing this method | Summary of the evidence |
|---|---|---|---|
| | | | blood collection followed by volumetric assessment, with a mean difference in recorded blood loss of 58.6ml (28). One study of 228 women with PPH following vaginal delivery found weighing blood loss compared to Hgb drop (of 10%) had a sensitivity of <75% and a specificity of 97%). These findings were modelled at hypothetical high prevalence PPH settings (15%, 30%), where the Positive Predictive Value was >86% (29). |
| **METHODS OF DETECTING HEMODYNAMIC INSTABILITY SECONDARY TO PPH DURING AND AFTER CAESAREAN BIRTH** | | | |
| *Clinical signs of haemodynamic instability* | • N/A | 6 Guidelines:<br>(2, 3, 5, 10, 12, 14)<br>3 Systematic reviews:<br>(19, 20, 21)<br>Additional relevant references:<br>(30, 31, 32, 33, 34, 35, 36) | Signs of haemodynamic instability reported in guidelines included changes in blood pressure, heart rate, pulse oximetry, urine output, or general status (faintness/dizziness, nausea, thirst, altered level of consciousness, pallor, sweating, poor capillary refill, and cold extremities. The guidelines proposed considering clinical signs and symptoms of haemodynamic instability in combination with other methods (such as volumetric and gravimetric methods for blood loss assessment) for the detection of PPH.(2, 3, 5, 10, 12, 14). Some of these guidelines presented tables correlating clinical signs with blood loss and the degree of shock. However, most of them warned that many clinical signs and symptoms do not occur until the blood loss reaches very high levels due to the physiological increase in circulating blood volume during pregnancy. Some other guidelines (6, 12, 13, 14) explicitly proposed including clinical signs and symptoms of haemodynamic instability or the use of the shock index specifically in assessing PPH severity. None of the trials included in the systematic reviews used this method to detect PPH. The Shock Index (SI) as a predictor of several maternal outcomes has been evaluated in the context of PPH research, including both vaginal and caesarean birth. SI performance was usually reported using the Area under the Curve (AUC) parameter (with estimates between 0.7 - 0.8 in most studies). According to the cut-off value of the SI chosen and the specific outcome analysed, SI sensitivity may range from 30% to 90%(30, 31, 32, 33, 34, 35). A recent stepped-wedge cluster randomized trial showed insufficient evidence to suggest that a significant benefit or harm could be attributed using an automated SI device (in low-resource settings) on a composite outcome of maternal deaths, eclampsia, or emergency (36). However, the rate of emergency hysterectomy was significantly reduced. |
| *Visual charts and early warning scores (EWS)* | • N/A | 5 Guidelines:<br>(2, 4, 6, 12, 13)<br><br>0 Systematic reviews:<br>None reference this method.<br><br>Additional relevant references:<br>(37) | A few guidelines recommended visual charts and early warning scores to alert caregivers to abnormal trends in haemodynamic measurements. However, these seem to be recommended for follow-up monitoring of diagnosed PPH cases rather than for diagnosis of PPH. None of the trials included in the systematic reviews used this method to detect PPH. A systematic literature review of 17 published obstetric EWS reported that they had very high median sensitivity (89%) and specificity (85%) but low median positive predictive values (41%) for predicting morbidity or ICU admission. Obstetric EWS had high accuracy in predicting death (AUROC >0.80) among critically ill obstetric women (37). |

**Table S3.** Synthesis of evidence on thresholds for triggering action on PPH during and after caesarean birth

| Thresholds | Guidelines recommending this threshold | Applicable mode of birth (if thresholds differ by mode of birth) | Comments |
|---|---|---|---|
| **ONE-STEP APPROACH: THRESHOLDS TRIGGER FULL RESPONSE PROTOCOL** | | | |
| *Blood loss at least 1000 mL OR signs of haemodynamic instability* | 1. (10)<br>2. (11)<br>3. (12)<br>4. (9) | Three guidelines proposed this threshold as being specific for caesarean birth(9, 10, 12). | In these guidelines the same criteria proposed as the definition of PPH were also used as the threshold for initiating treatment. |
| *Blood loss at least 500 mL* | 1. (6) | This guideline stated that this threshold applied regardless of the mode of birth. | The guideline noted that clinical signs and symptoms of hypovolaemia should be included in the assessment of PPH severity. However, clinical signs of hypovolaemia are misleading in pregnancy due to plasma volume expansion and might not become evident until blood losses reach 1,000-1,500 mL in healthy women. Thus, the blood loss thresholds should depend on the woman's clinical condition and local resources. In this guideline the same criteria proposed for the definition of PPH were also used as the threshold for initiating treatment. |
| *Blood loss at least 500 mL OR Signs of haemodynamic instability* | 1. (3) | This guideline stated that this threshold applied regardless of the mode of birth. | This guideline noted that the bleeding rate, PPH etiology, and clinical context should be considered. Further, for caesarean births, thresholds could be set at a higher blood loss if clinically tolerated. In this guideline the same criteria proposed for the definition of PPH were also used as the threshold for initiating treatment. |
| *Blood loss at least 1000 mL* | 1. (7) | This guideline stated that this threshold is specific for caesarean birth. | In this guideline the same criteria proposed for the definition of PPH were also used as the threshold for initiating treatment. |
| *Any excessive bleeding with signs of hemodynamic instability* | 1. | This guideline stated that this threshold is applicable to all births. | In this guideline the same criteria proposed for the definition of PPH were also used as the threshold for initiating treatment. |
| **TWO/STEP APPROACH: THRESHOLDS TRIGGER DIFFERENTIAL ACTIONS** | | | |
| *Lower threshold: Blood loss at least **500 mL** without clinical shock; Higher threshold: Blood loss at least **1000 ml,** continued bleeding, OR clinical shock* | 1. (2)<br>2. (4)<br>3. (5)<br>4. (13) | Guidelines did not specify a difference in threshold by mode of birth. | According to these guidelines, blood loss of **500–1000 ml** (minor PPH) without clinical shock should trigger: close monitoring, laboratory tests, and the use of crystalloid infusion [2] ; prompt basic measures (close monitoring, intravenous access, full blood count, group, and screen, insert urinary catheter) to facilitate resuscitation [4] ; enhanced surveillance and early interventions as needed[13] . Blood loss ≥ **1000 ml** and continued bleeding or clinical shock should trigger a full protocol to achieve resuscitation and haemostasis. [2, 4, 13]Three guidelines aligned their proposed lower thresholds with their proposed PPH definitions. [2, 4, 5] A single guideline aligned its proposed higher threshold with its PPH definition. [13] |

| Thresholds | Guidelines recommending this threshold | Applicable mode of birth (if thresholds differ by mode of birth) | Comments |
|---|---|---|---|
| *Lower threshold: Blood loss at least 1000 mL; Higher threshold: Blood loss of at least 2000 mL OR SI of ≥1.0* | 1. (8) | Guideline provided thresholds specific for CB | According to this guideline, blood loss of **1000 mL** should trigger suspicion of PPH and initiation of treatment. Blood loss of *at least* **2000 mL or SI of ≥1.0** should trigger: initiation of IV catheter with a large gauge and replacement of a sufficient volume of fluid; consideration of blood transfusion and the transportation of the patient to a secondary or tertiary hospital; monitoring of blood pressure, pulse rate, bleeding amount, urine output and SpO2. This guideline used the same criteria as thresholds for action as were used for proposed PPH definition. |

*Four guidelines (1, 15, 16, 17)did not mention any thresholds for initial assessment or to trigger a full protocol. Note CB= caesarean birth

**Table S4.** Medical Interventions, manoeuvres, and procedures for PPH recommended by WHO compared to recommendations in other PPH guidelines and systematic reviews

| Method | WHO recommendation (PPH 2012,TXA 2017, Carbetocin 2018, and UBT 2021) (1, 9, 38, 39) | Other Reviewed Guidelines | Systematic Reviews |
|---|---|---|---|
| **MEDICAL INTERVENTIONS** | | | |
| **Uterotonics** | | | |
| *Oxytocin* | Intravenous oxytocin was the recommended first-line treatment for PPH, including among those who have already received oxytocin for prevention of PPH (no dosing information included; 10 IU IM or IV was the recommended dosing and route of administration provided for prophylactic use). | Recommended as first line drug ((2, 3, 4, 5, 6, 7, 10, 12, 13, 14, 17)<br>Two different dosing regimens discussed:<br>• Intravenous oxytocin 5 IU (slow IV injection over 2 minutes). May repeat dose once.<br>• Intravenous infusion 5-10 IU per hour (20-40 IU in 500 ml saline over 4 hours). | Not described. |
| *Carbetocin* | Not described in the 2012 WHO treatment guidelines. The 2018 WHO prevention guidelines recommended prophylactic carbetocin (100 µg, IM/IV) for all births when cost was comparable to other effective uterotonics, but did not recommend the use of carbetocin as a treatment for PPH. | Recommended as a first line drug (7, 14)or as a second-line drug (10) for treatment. | Not described. |
| *Ergometrine* | Recommended if IV oxytocin unavailable, or bleeding nonresponsive to oxytocin. No dosing amount provided. Intravenous route of administration recommended. | Intravenous or intramuscular routes recommended. Maximum dose of 1000 mcg. 250-500 mcg (IV slow, over 2 minutes, or IM), may repeat every 5 minutes (2, 4, 5, 12) | Ergometrine 200 mcg administered intramuscularly, followed by 250 mcg IM carboprost if needed (21) |
| *Oxytocin-ergometrine fixed dose* | Recommended if IV oxytocin unavailable, or bleeding nonresponsive to oxytocin. No dosing amount or route of administration recommended. ` | Not described. | Syntometrine® (ergometrine 500 mcg plus oxytocin 5 IU) administered intramuscularly plus oxytocin 10 IU administered by an intravenous infusion (21) |

| Method | WHO recommendation (PPH 2012,TXA 2017, Carbetocin 2018, and UBT 2021) (1, 9, 38, 39) | Other Reviewed Guidelines | Systematic Reviews |
|---|---|---|---|
| *Prostaglandin (including sublingual misoprostol, 800 mcg)* | Recommended if IV oxytocin unavailable, or bleeding nonresponsive to oxytocin. No dosing amount or route of administration information provided other than for misoprostol (recommended 800 mcg, administered sublingually). Sublingual misoprostol particularly recommended in settings where IV oxytocin unavailable and IM oxytocin used for prophylaxis. | Carboprost was recommended either: as a general second line drug (12), specifically for use after oxytocin/ergometrine (2), or recommended generally with no specific order cited (5). Most reviewed guidelines recommended a dose of 250 mcg IM, which could be repeated every 15 minutes, up to a maximum dose of 2000 mcg (2, 3, 4, 5, 6, 10, 11, 12, 13, 14). An alternative dosing regimen of 500 mcg intramyometrial route was cited in 6 guidelines (4, 5, 11, 12, 13, 14)Misoprostol was recommended either: as first line drug (5), when other first-line drugs unavailable or contraindicated (12), after oxytocin/ergometrine (2), or if carbroprost contraindicated (4). The recommended dosing regimen was a single dose of 600-1000 mcg, by oral, sublingual, or rectal route (2, 3, 4, 5, 6, 7, 10, 11, 12, 13, 14). Sulprostone IV route, 500 mcg administered over 1 hour (3, 6, 7) | Carboprost 250 mcg administered intramuscularly, followed by 250 mcg IM ergometrine if needed – considered a second line drug (21).<br>Misoprostol 800 mcg (4 tablets of 200 mcg) administered rectally. Considered a first line drug. Data drawn from seven trials, only one of which included some women with caesarean birth (21) |
| *Tranexamic acid with standard care* | The 2017 WHO guidelines on tranexamic acid recommended a fixed dose of 1 g (100 mg/mL) administered intravenously at 1 mL per minute (i.e., administered over 10 minutes), with a second 1 g IV dose if bleeding continued after 30 minutes OR restarted within 24 hours of completing the first dose. | Intravenous administration of 1 g over 10 minutes. A second dose may be administered after 30 minutes if bleeding persists (2, 3, 4, 5, 6, 7, 11, 12) | A fixed dose of 1g (100 mg/mL) intravenously at 1 ml per minute, within 3 hours of the time of diagnosis (if unknown, time of birth); a second dose of 1g given if needed 30 minutes from the first dose. Considered a first-line treatment (40). |
| **MANOEUVRES AND OTHER PROCEDURES** | | | |
| **Mechanical interventions** | | | |
| *Uterine massage* | Rubbing of the uterus achieved through manual massage of the abdomen, typically sustained until bleeding ceases or the uterus contracts. (initial rubbing of uterus and expression of clots NOT considered therapeutic uterine massage).<br>Recommended (low cost and relative safety of uterine massage considered in this recommendation).<br>Note: This recommendation was developed considering vaginal birth. | Intervention recommended in other 10 guidelines (2, 3, 4, 5, 7, 10, 11, 12, 14) | Not described. |

| Method | WHO recommendation (PPH 2012,TXA 2017, Carbetocin 2018, and UBT 2021) (1, 9, 38, 39) | Other Reviewed Guidelines | Systematic Reviews |
|---|---|---|---|
| *Intrauterine balloon tamponade* | The procedure entails insertion of a deflated/uninflated balloon into the uterine cavity and then inflating it to achieve a tamponade effect. Uterine balloon tamponade was recommended for the treatment of postpartum haemorrhage due to uterine atony after vaginal birth in women who did not respond to standard first-line treatment, provided all required resources for of PPH are available and routinely implemented (39). Only two studies included in the evidence supporting the recommendations included caesarean deliveries, both of which evaluated the effect of UBT in cases of placenta praevia or traumatic bleeding. One study (41)suggests that the use of the Bakri balloon could be more effective than haemostatic sutures in, and the second (42)suggests benefits associated to the use of the Bakri balloon held in place with a traction stitch versus Bakri balloon without traction stitch. | Guidelines described the urological Rusch balloon (left over 4-6 hrs), the Bakri SOS tamponade balloon catheter, the Sengstaken-Blakemore esophageal catheter, the Foley catheter, the polyurethane Ebb double balloon (vaginal and uterine), and the silicone BT-Cath tamponade balloon (2, 4, 6, 7, 10, 11, 13, 14) | One trial (50 women) compared Bakri Balloon with and without traction stitch; another trial (13 women) compared Bakri balloon to compressive suturing to the lower segment of the uterus. Not specified whether a first-line, second-line, or temporising treatment (20) |
| *Uterine packing* | Not recommended for PPH due to uterine atony. | One guideline (13) did not recommend the use of uterine packing while two guidelines recommended its use as a temporizing measure (14) or for unresponsive PPH (11). | Not described. |
| **Uterine-sparing surgical interventions and procedures** | | | |
| *Compressive sutures* | Compression suturing that runs through the full thickness of both uterine walls. When tied, the suture allows tight compression of the uterine walls and stops the bleeding. No specific suturing technique (e.g., B-Lynch, Hayman, Pereira). Recommended as first-line surgical intervention. | The B-Lynch technique was the most common uterine compression technique for atony (2, 3, 4, 5, 7, 10, 11, 12, 13, 14); however, other techniques, such as Cho and Hayman, were also recommended and described (10, 11). | One trial (160 women) compared the standard B-Lynch suture to a modified B-Lynch suture. Considered a second-line treatment (20). |
| *Devascularisation / Artery ligation* | Vascular flow to the uterus can be interrupted by uterine devascularization, ligation of the uterine or internal iliac arteries Recommended only if all available conservative measures (uterotonics, uterine massage, balloon tamponade) have failed. | If compression sutures are unsuccessful, bilateral uterine artery ligation, bilateral utero-ovarian artery ligation or -If expertise available- bilateral internal iliac artery ligation must be considered (2, 3, 4, 5, 6, 7, 10, 11, 12, 14). A common first approach is bilateral uterine artery ligation (O'Leary sutures) (11). | One trial (23 women) compared uterine artery embolization to surgical devascularization plus B-Lynch compression sutures. Not specified wither first-response, second-line, or temporizing treatment (20). |

| Method | WHO recommendation (PPH 2012,TXA 2017, Carbetocin 2018, and UBT 2021) (1, 9, 38, 39) | Other Reviewed Guidelines | Systematic Reviews |
|---|---|---|---|
| *Uterine artery embolisation (UAE)* | If other measures have failed and if the necessary resources were available, the use of uterine artery embolization was recommended as a treatment for PPH due to uterine atony. | Twelve guidelines recommended UAE (1, 2, 3, 4, 5, 6, 7, 10, 11, 12, 13, 14).<br>Only one describes the intervention in greater detail(3) | One trial (23 women) compared uterine artery embolization to surgical devascularization plus B-Lynch compression sutures. Not specified wither first-line, second-line, or temporizing treatment (20). |
| **Temporising** | | | |
| *External aortic compression* | Recommended as a temporizing measure until appropriate care is available, in PPH due to uterine atony. | In addition to WHO guidelines, seven guidelines recommended external aortic compression (Queensland, (2, 4, 6, 7, 10, 12, 14) | Not described in Systematic Reviews. |
| *Non-pneumatic anti-shock garment* | Recommended as a temporizing measure until appropriate care is available. | Recommended in other three guidelines (6, 10, 13). | One systematic review did not find a reduction in maternal mortality associated to NASG in the one cluster-RCT (880 women) included, However, 5 comparative studies (pre-intervention to intervention) included in this review (2330 women) suggested a clinically important reduction in maternal mortality and severe maternal morbidity(43). No effect was observed on the use of blood products. There were no safety issues in all the trials (43). |

*Pileggi-Castro 2015 (43) is a systematic review on the non-pneumatic anti-shock garment as a treatment for severe PPH. This review was not captured as part of the overview review because the word “cesarean”/”caesarean” did not appear in the text. After consultation with Prof. Suellen Miller, who participated in each of the primary studies, confirmed that women with caesarean section were included in the original trials, data from this systematic review was added to the report and is reflected in the following tables.

**Table S5**. First and second round ratings and agreement for early detection methods for intraoperative and postoperative CB-PPH

| *Blood loss measurement and other PPH detection methods* | How would you rate each of the methods below for early detection of PPH considering… | | | | | | | |
|---|---|---|---|---|---|---|---|---|
| | the usefulness in managing patients? | feasibility in all settings performing CB? | its acceptability to key stakeholders? | the resources required?* | the usefulness in managing patients? | feasibility in all settings performing CB? | its acceptability to key stakeholders? | the resources required?* |
| | 1=Not at all useful; 9=extremely useful | 1=Hardly feasible; 9=Highly feasible | 1=Not at all useful; 9=extremely useful | 1= very small; 9=very large | 1=Not at all useful; 9=extremely useful | 1=Hardly feasible; 9=Highly feasible | 1=Not at all useful; 9=extremely useful | 1= very small; 9=very large |
| | FIRST ROUND Median (DI) | | | | SECOND ROUND Median (DI) | | | |
| **INTRAOPERATIVE** | | | | | | | | |
| Volumetric/gravimetric + clinical signs of haemodynamic instability | **8.0 (-0.93)** | 6.0 (2.35) | NA | NA | NA | **8.0 (-3.08)** | NA | NA |
| Volumetric | **8.0 (-0.71)** | 6.5 (3.50) | 7.0 (2.35) | 6.5 (1.85) | NA | 6.5 (10.00) | 7.0 (10.0) | 7.0 (2.35) |
| Clinical signs of haemodynamic instability | 6.5 (8.31) | **8.0 (-1.94)** | **8.0 (-0.22)** | **3.5 (0.58)** | **7.0 (-0.71)** | NA | NA | NA |
| Volumetric + gravimetric | **7.0 (-1.26)** | **5.0 (0.94)** | NA | NA | NA | NA | NA | NA |
| Clinical judgement such as rate of flow and duration | **5.0 (0.92)** | **7.0 (-21.7)** | 6.0 (2.35) | **2.0 (0.49)** | NA | NA | 7.0 (10.0) | NA |
| Visual charts and early warning scores (EWS) | **5.0 (0.88)** | 6.0 (2.09) | 6.0 (1.96) | 5.0 (1.70) | NA | 7.0 (10.0) | 6.5 (10.0) | 6.0 (1.74) |
| Visual estimation of blood loss | **4.5 (0.97)** | **8.5 (-1.81)** | 7.0 (30.0) | **1.0 (0.13)** | NA | NA | **7.0 (-1.94)** | NA |
| Gravimetric | 6.0 (2.35) | **4.5 (0.91)** | **5.0 (0.85)** | 6.0 (2.09) | 7.0 (10.00) | NA | NA | 7.0 (10.00) |
| Visual estimation + visual charts/EWS | 6.0 (1.48) | **7.0 (-0.71)** | NA | NA | 6.5 (2.35) | NA | NA | NA |
| **POSTOPERATIVE** | | | | | | | | |
| Clinical signs of haemodynamic instability | **8.0 (-3.08)** | **8.0 (-1.27)** | **8.0 (-1.94)** | **3.5 (0.58)** | NA | NA | NA | NA |
| Volumetric/gravimetric + clinical signs of haemodynamic instability | **7.0 (-0.93)** | 6.0 (1.74) | NA | NA | NA | 7.0 (30.00) | NA | NA |
| Clinical judgement such as rate of flow and duration | **5.0 (0.41)** | 6.0 (2.35) | 6.0 (2.35) | **2.0 (0.49)** | NA | 6.0 (2.35) | 7.0 (10.00) | NA |
| Gravimetric | 5.5 (2.35) | **3.0 (0.65)** | **5.0 (0.85)** | 6.0 (2.09) | **5.0 (0.63)** | NA | NA | 7.0 (10.00) |
| Volumetric | 5.0 (2.09) | **4.5 (0.58)** | 7.0 (2.35) | 6.5 (1.85) | **5.0 (0.63)** | NA | 7.0 (10.00) | 7.0 (2.35) |
| Visual estimation of blood loss | **4.0 (0.52)** | **7.0 (-3.08)** | 7.0 (30.0) | **1.0 (0.13)** | NA | NA | **7.0 (-1.94)** | NA |
| Visual charts and early warning scores (EWS) | 6.0 (2.09) | 7.0 (2.35) | 6.0 (1.96) | 5.0 (1.70) | 7.0 (10.0) | 7.0 (10.00) | 6.5 (10.00) | 6.0 (1.74) |
| Visual estimation + visual charts/EWS | 6.0 (4.96) | **7.0 (-3.08)** | NA | NA | 7.0 (10.0) | NA | NA | NA |
| Volumetric + gravimetric | 7.0 (8.31) | 4.0 (1.04) | NA | NA | 7.0 (10.0) | **5.0 (0.85)** | NA | NA |

Note: A DI < 1 represented agreement, while a DI ≥ 1 indicated disagreement. Results in which agreement is reached are highlighted in bold.NA= Not applicable given that this combination of methods was not rated in the first round for acceptability to key stakeholders and the estimate of resources required, or because agreement was obtained in the first round. * The measurement scale for this criterion is the same as for the other criteria (from 1 to 9). However, unlike the other criteria, low values have a positive interpretation (few resources required) while high values have a negative interpretation (substantial resources required).

**Table S6.** First round ratings and agreement on threshold to initiate treatment for intraoperative CB PPH

| *Thresholds for triggering action* | **How would you rate each of the following thresholds for managing PPH during CB considering…** | | |
|---|---|---|---|
| | **the accuracy of each threshold?** 1=Hardly accurate; 9=Highly accurate | **its feasibility to be used in all settings?** 1=Hardly feasible; 9=Highly feasible | **its acceptability to key stakeholders?** 1=Hardly accepted; 9=Highly accepted |
| **INTRAOPERATIVE** | | | |
| **One-step approach (trigger full response protocol)** | | | |
| At least 1000 mL blood loss OR signs of haemodynamic instability, whichever comes first | **9 (-0.34)** | **8 (-0.34)** | **8 (-0.54)** |
| At least 1000 ml (blood loss alone, regardless of signs of haemodynamic instability) | **7 (-0.71)** | **8 (-0.71)** | **7 (-2.30)** |
| Haemodynamic instability alone, regardless of volume of blood loss | **7 (-3.08)** | **7 (-0.71)** | **7 (-3.08)** |
| At least 500 mL blood loss OR signs of haemodynamic instability, whichever comes first | 7 (2.30) | **7 (-4.00)** | 6.5 (8.31) |
| At least 500 mL (blood loss alone, regardless of signs of haemodynamic instability) | **5 (0.41)** | 6.5 (2.14) | **5 (0.88)** |
| **Two-step approach (Lower threshold triggers further assessment, preparedness, and close monitoring; Higher threshold triggers treatment initiation)** | | | |
| Lower threshold of blood loss at least 500 mL (blood loss alone, regardless of signs of haemodynamic instability), and higher threshold of blood loss at least 1000 mL blood loss OR signs of haemodynamic instability, whichever comes first | **8 (-0.71)** | **8 (-0.71)** | **7 (-0.71)** |
| Lower threshold of blood loss at least 1000 ml (blood loss alone, regardless of signs of haemodynamic instability), and higher threshold of blood loss at least 2000 mL blood loss OR signs of haemodynamic instability whichever comes first | 6.5 (4.96) | **8 (-4.23)** | 7 (30.00) |
| **POSTOPERATIVE** | | | |
| **One-step approach (trigger full response protocol** | | | |
| At least 1000 mL blood loss OR signs of haemodynamic instability, whichever comes first | **8.5 (-0.34)** | **8 (-2.14)** | **8 (-0.88)** |
| At least 1000 ml (blood loss alone, regardless of signs of haemodynamic instability) | **7 (-0.71)** | **8 (-0.71)** | **7 (-1.27)** |
| Haemodynamic instability alone, regardless of volume of blood loss | **7 (-3.08)** | **7 (-4.00)** | 7 (10.00) |
| At least 500 mL blood loss OR signs of haemodynamic instability, whichever comes first | 7 (8.31) | **7 (-16.8)** | 6.5 (30.00) |
| At least 500 mL (blood loss alone, regardless of signs of haemodynamic instability) | 5 (1.47) | 6 (2.09) | 5 (1.18) |
| **Two-step approach (Lower threshold triggers further assessment, preparedness, and close monitoring; Higher threshold triggers treatment initiation)** | | | |
| Lower threshold of blood loss at least 500 mL (blood loss alone, regardless of signs of haemodynamic instability), and higher threshold of blood loss at least 1000 mL blood loss OR signs of haemodynamic instability, whichever comes first | **8 (-0.71)** | **8 (-0.71)** | **7.5 (-0.71)** |

| | | | |
|---|---|---|---|
| Lower threshold of blood loss at least 1000 ml (blood loss alone, regardless of signs of haemodynamic instability), and higher threshold of blood loss at least 2000 mL blood loss OR signs of haemodynamic instability whichever comes first | 6.5 (4.41) | 6 (2.35) | 6 (1.96) |

Note: A DI < 1 represented agreement, while a DI ≥ 1 indicated disagreement. Results in which agreement is reached are highlighted in bold. CB= caesarean birth.

**Table S7.** First round ratings and agreement for first response interventions for managing intraoperative and postoperative CB-PPH

| *First response interventions for managing PPH during and after CB* | **How would you rate each intervention for first response management of CB-PPH considering…** | | | | |
|---|---|---|---|---|---|
| | **the balance of effects?** 1=Weighted towards undesirable effects; 9=Weighted towards desirable effects | **the resources required? *** 1= very few; 9=very many | **its feasibility?** 1=Hardly feasible; 9=Highly feasible | **its acceptability to stakeholders?** 1=Hardly accepted; 9=Highly accepted | **equity?** 1=Likely to exacerbate inequities; 9=Highly likely to reduce inequities |
| | Median | | | | |
| ***INTRAOPERATIVE*** | | | | | |
| Oxytocin | **9 (-0.34)** | 3 (5.86) | **9 (-0.34)** | **9 (0.00)** | **9 (0.00)** |
| Carbetocin | **8 (-0.93)** | 5 (1.81) | **7.5 (-3.79)** | **8 (-0.65)** | 6 (6.55) |
| TXA | **8 (-0.34)** | **4 (0.76)** | **7 (-4.00)** | **8 (-0.93)** | **8 (-3.79)** |
| Compressive sutures | 7 (2.9) | 7 (4.64) | 6 (4.71) | 5.5 (1.76) | 6 (4.71) |
| Bimanual compression | 7 (10.15) | **3 (0.64)** | **7.5 (-3.08)** | 6.5 (12.8) | **8 (-2.56)** |
| Uterine massage | **7 (-15.2)** | 2 (1.17) | **8.5 (-0.65)** | **8 (-0.93)** | 8 (-0.34) |
| Oxytocin-ergometrine fixed dose | 6 (2.35) | 4 (1.61) | **7 (-3.08)** | **8 (-3.08)** | **8 (-2.19)** |
| Prostaglandin (including sublingual misoprostol) | 6 (1.37) | **3 (0.87)** | **8 (-3.79)** | **8 (-2.14)** | **8 (-0.93)** |
| Ergometrine | 6 (4.00) | **3 (0.91)** | **8 (-0.92)** | **7 (-0.92)** | **8 (-0.34)** |
| Non-pneumatic anti-shock garment | 5 (1.04) | **6 (0.78)** | **4 (0.52)** | 5 (1.08) | 5 (1.00) |
| External aortic compression | **5 (0.56)** | **2 (0.75)** | 6 (30.00) | **5 (0.78)** | 7 (30.00) |
| Intrauterine balloon tamponade | 4 (1.61) | 5 (2.14) | **4 (0.91)** | **5 (0.85)** | 6 (4.22) |
| ***POSTOPERATIVE*** | | | | | |
| Oxytocin | **9 (-0.34)** | 3 (5.86) | **9 (-0.34)** | **9 (0.00)** | **9 (0.00)** |
| TXA | **8 (-0.34)** | **4 (0.76)** | **7 (-13.00)** | **8 (-0.93)** | **8 (-6.78)** |
| Non-pneumatic anti-shock garment | 6 (1.35) | 6 (1.73) | **4 (0.52)** | **5 (0.99)** | 5 (1.00) |
| Carbetocin | **6 (-10.29)** | 5 (1.81) | **7.5 (-8.76)** | **8 (-0.92)** | 5.5 (4.22) |
| Oxytocin-ergometrine fixed dose | 6 (2.05) | 4 (1.61) | **7 (-3.08)** | **8 (-3.08)** | **8 (-1.54)** |
| Uterine massage | 6 (2.35) | 3 (1.17) | **8 (-0.93)** | **8 (-1.68)** | **8 (-3.79)** |
| Ergometrine | 6 (4.00) | **3 (0.97)** | **8 (-0.71)** | **7.5 (-2.19)** | **8 (-0.34)** |
| Prostaglandin (including sublingual misoprostol) | **6 (0.89)** | 3 (1.35) | **8 (-1.53)** | **7.5 (-2.14)** | **8 (-0.93)** |
| Bimanual compression | **5 (0.63)** | **3 (0.41)** | 6 (16.57) | 6 (1.73) | **8 (-23.00)** |
| External aortic compression | **5 (0.68)** | **2 (0.68)** | 6 (2.25) | **5.5 (0.52)** | 7 (5.6) |
| Intrauterine balloon tamponade | 4 (1.64) | 5 (2.14) | **4.5 (0.91)** | **5 (0.85)** | 4.5 (4.22) |
| Compressive sutures | **2 (0.89)** | 6 (2.84) | **4 (0.99)** | **3 (0.91)** | 4.5 (1.59) |

Note: A DI < 1 represented agreement, while a DI ≥ 1 indicated disagreement. Results in which agreement is reached are highlighted in bold. CB= caesarean birth. * The measurement scale for this criterion is the same as for the other criteria (from 1 to 9). However, unlike the other criteria, low values have a positive interpretation (few resources required) while high values have a negative interpretation (substantial resources required)

**Table S8.** Second round ratings and agreement for first response interventions for managing intraoperative and postoperative CB-PPH

| **Intraoperative** | Median (RAND DI) |
|---|---|
| Examine and rapidly initiate cause-specific first response (e.g., if trauma: rapid surgical haemostasis; if atony/placental cause: uterotonics and uterine massage) | **9 (-0.34)** |
| TXA for all women with PPH during CB regardless of aetiology | **8 (-0.43)** |
| Plasma expansion with crystalloids or all women with PPH during CB regardless of aetiology | **7.5 (-1.94)** |
| Uterotonics for all women with PPH during CB regardless of aetiology | **7 (-0.71)** |
| **Postoperative** | |
| Examine and rapidly initiate cause-specific first response (e.g., if trauma: rapid surgical haemostasis; if atony/placental cause: uterotonics and uterine massage) | **9 (-0.34)** |
| TXA for all women with PPH during CB regardless of aetiology | **8 (0.00)** |
| Plasma expansion with crystalloids or all women with PPH during CB regardless of aetiology | **7.5 (-0.71)** |
| Uterotonics for all women with PPH during CB regardless of aetiology | **7 (-0.71)** |

Note: A DI < 1 represented agreement, while a DI ≥ 1 indicated disagreement. Results in which agreement is reached are highlighted in bold. CB= caesarean birth.

**Supplementary Figures**

**Figure S1.** Interpretation of DI and RAND/UCLA Appropriateness scale

| DI (Disagreement index) | Experts' median rating | | | | | | | | |
|---|---|---|---|---|---|---|---|---|---|
| | 1 | 2 | 3 | 4 | 5 | 6 | 7 | 8 | 9 |
| | Bottom third (1-3) | | | Intermediate third (4-6) | | | Top third (1-3) | | |
| <1 (Agreement) | Inappropriate | | | Uncertain | | | Appropriate | | |
| ≥1 (Disagreement) | Uncertain | | | | | | | | |

**Figure S2.** PRISMA Flowchart

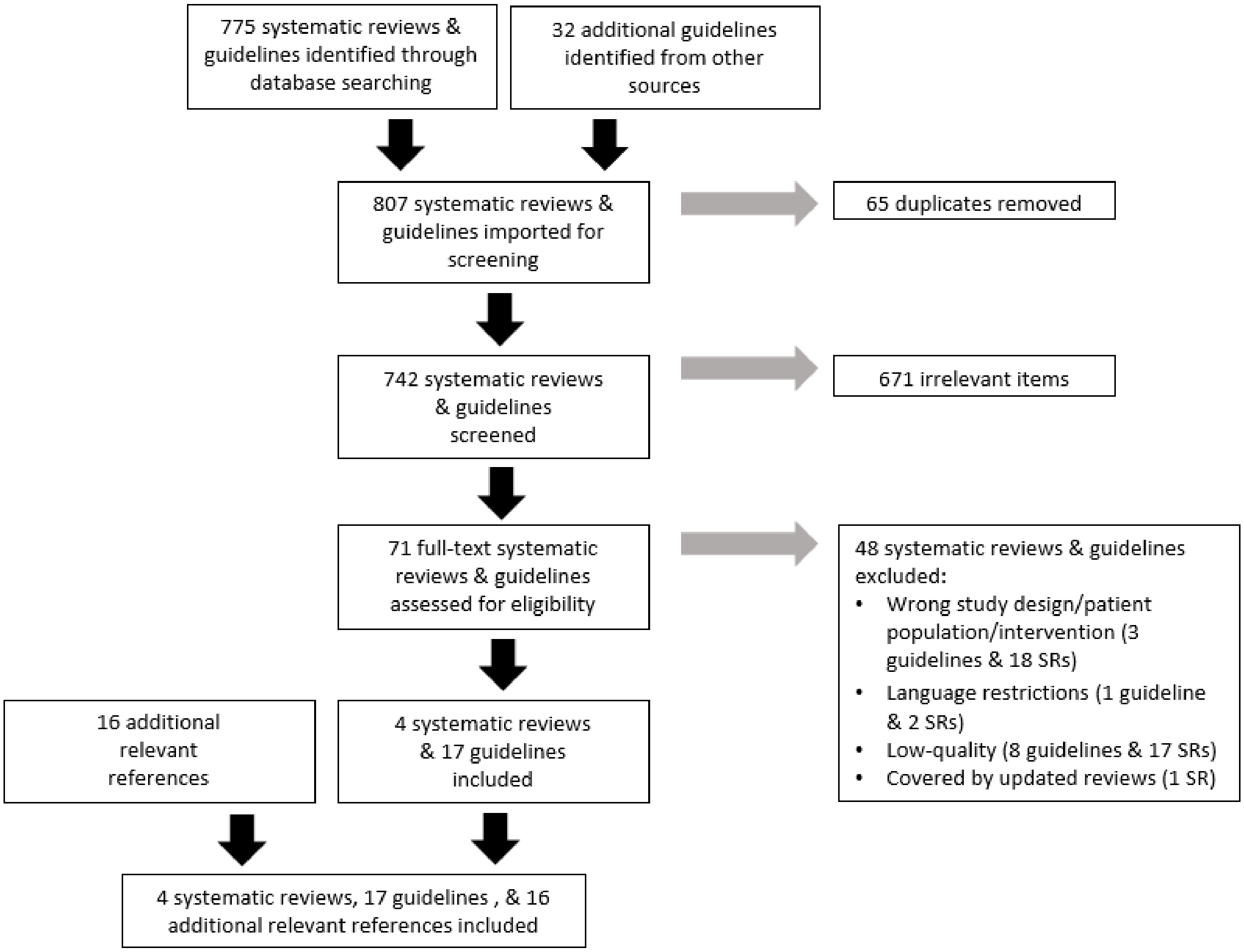

**Figure S3.** Second round ranking of one-step thresholds to initiate treatment for intraoperative and postoperative CB PPH

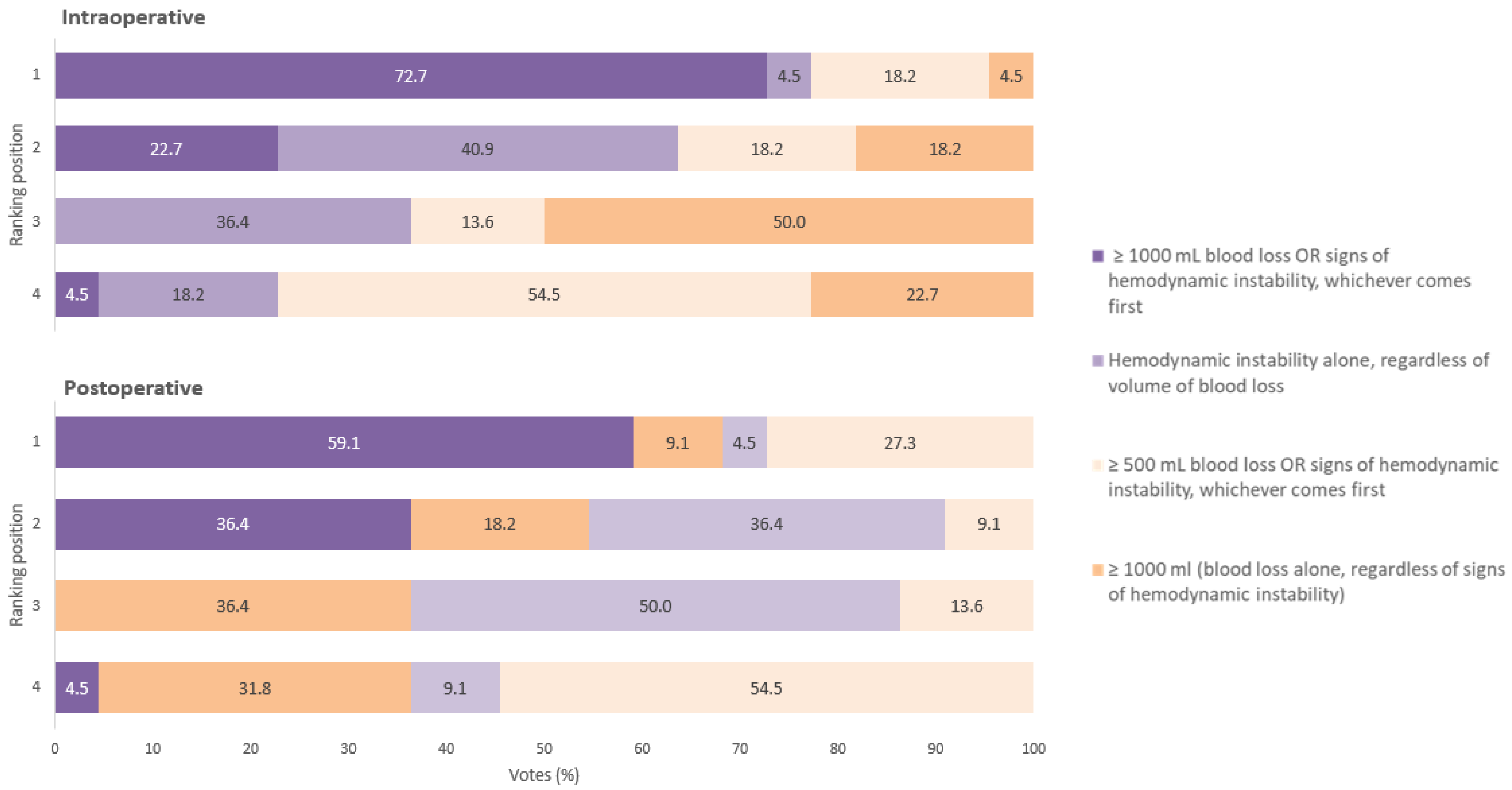

Note: CB= caesarean birth

**Supplementary Files**

**Supplementary File S1.** Search strategy

**(Postpartum Hemorrhage[Mesh] OR** Hemorrhag*[tiab] OR Haemorrhag*[tiab] OR PPH*[tiab] OR **Blood Specimen Collection[Mesh]** OR Blood Loss[tiab] OR "Loss of Blood"[tiab] OR Bleeding[tiab] OR Blood Specimen[tiab] OR Blood Collection[tiab]) AND **(Cesarean Section[Mesh**] OR C-Section*[tiab] OR Cesaerea*[tiab] OR Cesarea*[tiab] OR Cesaria*[tiab] OR Caesaria*[tiab] OR Caesarea*[tiab]) **AND (Systematic Review[sb] OR** Systematic Review[tiab] OR **Meta-Analysis[pt]** OR Meta-Analys*[tiab] OR **"Cochrane Database Syst Rev"[ta]** OR Metaanalysis[tiab] OR Metanalysis[tiab] OR Overview[ti] OR (Review[ti] AND Literature[ti]) OR (MEDLINE[tiab] AND Cochrane[tiab]) OR **Guideline[pt]** OR **Practice Guideline[pt]** OR Guideline*[ti] OR Guide Line*[tiab] OR Consensus[tiab] OR Recommendation*[ti])

**(MH Postpartum Hemorrhage OR** Hemorrhag$ OR Hemorragia OR PPH$) AND **(MH Cesarean Section** OR C-Section$ OR Cesaerea$ OR Cesarea$ OR Cesaria$ OR Caesaria$ OR Caesarea$)

**Supplementary File S2.** In-person meeting agenda

### WHO Technical Consultation on early detection and first response to PPH during and after caesarean birth

**Meeting Agenda**

27-28 September 2022

Av. Appia 20, Geneva, Salle U2

Zoom: https://who.zoom.us/j/91683929578

(Meeting ID: 916 8392 9578; Password: &uqmG2dD)

**Introduction**

Thank you for your participation in the **WHO Technical Consultation on early detection and first response to PPH during and after caesarean birth**. This consultation is part of the ongoing EMOTIVE project (Early detection of Postpartum Haemorrhage and treatment using the WHO MOTIVE 'first response' bundle), coordinated by the University of Birmingham.

As a part of the EMOTIVE project, WHO and some EMOTIVE team members are conducting a three-stage modified Delphi process to generate consensus on the optimal approach for early detection and first-response treatment for postpartum haemorrhage occurring during (intraoperative) and after (postoperative) caesarean birth. The first two rounds of this consultation were informed by an overview of reviews of the literature and conducted via an anonymous online platform. This third, in-person round will serve to conclude the consultation.

**Experts' Meeting Objectives**

This meeting has two primary objectives:

To agree on an optimal CS-PPH detection strategy that would include blood loss measurement methods and thresholds for action during and after caesarean birth.

To develop a first response approach to manage PPH during and after caesarean birth. This objective includes both the selection of the strategy components and the identification of the optimal approach (bundle, algorithm, checklist, or a combination of them)

**Organization of the Meeting**

This meeting will be conducted in-person and via Zoom over the course of two (2) days. Please use this link to access the meeting virtually: https://who.zoom.us/j/91683929578 (Meeting ID: 916 8392 9578; Password: &uqmG2dD)

The first day of the meeting, we will present and discuss the results of the overview of reviews and first two rounds of the Delphi study. You will have the opportunity to ask questions regarding the findings. Later in the day, we will pose specific questions to the panel regarding issues raised by the results to-date. You will be asked to provide your opinion on each of these topics. Discussions will be focused first on the intraoperative period, and then the postoperative period. The objective is to present different points of view on methods for early detection of CS-PPH, thresholds for triggering action, and first-response treatments by the close of day 1.

The second day of the meeting, we will present proposed strategies for implementing early detection and first-response for CS-PPH in the intraoperative and postoperative periods. You will be asked to provide your opinions on the proposed strategies. Finally, you will be asked to rate the proposed strategies. The final results—including whether consensus was reached—and conclusion will be presented before the end of day 2.

**Dates and time**

| Date: 27-28 September 2022 | |
|---|---|
| *Day 1* | *Day 2* |
| Africa<br>• *Kenya — 10:00-18:05*<br>• *Nigeria — 08:00-16:05*<br>• *Republic of Congo —08:00-16:05*<br>• *South Africa — 09:00-17:05*<br><br>Americas<br>• *Argentina — 04:00-12:05*<br>• *Uruguay — 04:00-12:05*<br><br>Europe<br>• *Denmark — 09:00-17:05*<br>• *France (Paris) — 09:00-17:05*<br>• *Switzerland — 09:00-17:05*<br>• *United Kingdom — 08:00-16:05*<br><br>South-East Asia<br>• *Australia (Melbourne) — 17:00-01:05*<br>• *Egypt — 09:00-17:05*<br>• *Vietnam — 14:00-22:05*<br><br>Western Pacific<br>• *India — 12:30-20:35*<br>• *Philippines —15:00-23:05*<br>• *Thailand — 14:00-22:05* | U. Africa<br>• *Kenya — 11:00-14:05*<br>• *Nigeria — 09:00-12:05*<br>• *Republic of Congo — 09:00-12:05*<br>• *South Africa — 10:00-13:05*<br><br>Americas<br>• *Argentina — 05:00-08:05*<br>• *Uruguay — 05:00-08:05*<br><br>Europe<br>• *Denmark — 09:00-12:05*<br>• *France — 10:00-13:05*<br>• *Switzerland — 10:00-13:05*<br>• *United Kingdom — 09:00-12:05*<br><br>South-East Asia<br>• *Australia — 18:00-21:05*<br>• *Egypt — 10:00-13:05*<br>• *Vietnam — 15:00-18:05*<br><br>Western Pacific<br>• *India — 13:30-16:35*<br>• *Philippines — 16:00-19:05*<br>• *Thailand — 15:00-18:05* |

#### *AGENDA*

| **DAY 1: 27 September 2022** | | **Presenter** |
|---|---|---|
| ***9:00 – 9:30***<br>*30 min* | <u>*Opening session*</u><br>— Welcome and introductions<br>— Meeting objectives<br>— Meeting logistics | Dr. Olufemi Oladapo<br>Dr. Ioannis Gallos<br>Dr. Fernando Althabe<br>Ms. Caitlin Williams |
| ***9:30 – 10:30***<br>*60 min* | <u>*Session 1*</u><br>— Brief review of the objectives and methods of the Delphi study<br>— Briefly summarize the state of the literature<br>— Findings from the first and second rounds of the Delphi study<br>— Questions | Ms. Verónica Pingray<br>Ms. Caitlin Williams<br>Ms. Verónica Pingray<br>Prof. Suellen Miller |
| ***10:30 – 10:45***<br>*15 min* | <u>*Coffee/Tea Break*</u> | |
| ***10:45 – 12:45***<br>*120 min* | <u>*Session 2*</u><br>— Briefly review summary of findings on **intraoperative** period<br>— Guided discussions by topic | Ms. Verónica Pingray<br>Prof. Suellen Miller |
| ***12:45 – 13:30***<br>*45 min* | <u>*Lunch*</u> | |
| ***13:30 – 14:15***<br>*45 min* | <u>*Continue Session 2*</u> | Prof. Suellen Miller |
| ***14:15 – 15:45***<br>*90 min* | <u>*Session 3*</u><br>— Briefly review summary of findings on **postoperative** period<br>— Guided discussions by topic (detection, thresholds, first-response treatments) | Ms. Verónica Pingray<br>Dr. Fernando Althabe |
| ***15:45 – 16:00***<br>*15 min* | <u>*Coffee/Tea Break*</u> | |
| ***16:00 – 17:00***<br>*60 min* | <u>*Continue Session 3*</u> | Dr. Fernando Althabe |
| ***17:00 – 17:05***<br>*5 min* | <u>*Closing DAY 1*</u> | Dr. Ioannis Gallos |
| **DAY 2: 28 September 2022** | | |
| ***10:00 – 10:15***<br>*15 min* | <u>*Welcome*</u><br>— Objective and procedures for Day 2 | Dr. Fernando Althabe |
| ***10:15 – 11:00***<br>*45 min* | <u>*Session 1*</u><br>— Presentation of a revised list of early detection and first-response treatments for intraoperative and postoperative PPH<br>— Final voting for the intraoperative and postoperative interventions and presentation of results | Prof. Suellen Miller<br>Ms. Caitlin Williams |
| ***11:00 – 11:15***<br>*15 min* | <u>*Coffee/Tea Break*</u> | |
| ***11:15 – 12:35***<br>*80 min* | <u>*Session 2*</u><br>— Organization of an implementation strategy<br>— Guided discussion of possible additional considerations | Dr. Fernando Althabe<br>Prof. Suellen Miller |
| ***12:35 – 13:00***<br>*25 min* | <u>*Session 3*</u><br>— Conclusion and next steps | Dr. Fernando Althabe |
| ***13:00 – 13:05***<br>*5 min* | <u>*Closing DAY 2*</u> | Dr. Arri Coomarasamy<br>Dr. Olufemi Oladapo |

**Supplementary File S3.** In-person meeting discussion question guide

### Early detection and first response to postpartum haemorrhage during and after caesarean birth A Modified-Delphi Study

**Pending controversies or disagreements issues to discuss**

27-28 September 2022

**DAY 1: 27 September 2022**

**Definitions**:
The experts agreed that the same definition of PPH should be used for both vaginal and caesarean births (same regardless of the mode of birth). Currently the WHO defines PPH as blood loss at least 500 mL within 24 hours after birth.

**Draft proposed list of intraoperative interventions**

**Early detection of PPH and thresholds for triggering first-response management**

- Aspirated blood volume (+ weighted pads, sponges, gauzes, etc. if feasible) at least 1000 mL, OR
- Haemodynamic instability (blood pressure, heart rate, oximetry) with any blood loss volume

*# Register the final amount of intraoperative blood loss and hand over this information to recovery area.*

**First-response treatment**

- TXA (1g in 10 mL IV over 10 min) for all women
- Examine and rapidly initiate cause-specific first response:
  - If trauma: Rapid haemostasis: hysterorraphy, tears, wound.
  - If atony/placental cause: uterotonics and intra-abdominal uterine massage or exteriorize the uterus and massage
- Uterotonics for all women
- IV fluids with crystalloids for all women

**Pending issues**

**Thresholds**

**Background:** The experts agreed that blood loss of at least 1000 mL should be used as a threshold to trigger first-response management.

| | % | Ranking |
|---|---|---|
| **Intraoperative** | | |
| ≥1000 mL blood loss OR signs of hemodynamic instability, whichever comes first | 72.7 | 1 |
| Hemodynamic instability alone, regardless of volume of blood loss | 40.9 | 2 |
| ≥1000 ml (blood loss alone, regardless of signs of hemodynamic instability) | 50.0 | 3 |
| ≥500 mL blood loss OR signs of hemodynamic instability, whichever comes first | 50.0 | 4 |
| **Postoperative** | | |
| ≥1000 mL blood loss OR signs of hemodynamic instability, whichever comes first | 59.1 | 1 |
| Hemodynamic instability alone, regardless of volume of blood loss | 36.4 | 2 |
| ≥1000 ml (blood loss alone, regardless of signs of hemodynamic instability) | 36.7 | 3 |
| ≥500 mL blood loss OR signs of hemodynamic instability, whichever comes first | 50.0 | 4 |

We believe is important to discuss the overall implications of the different thresholds on clinical practice, in the context of the total blood that women lose during and after CS. At the meeting we will present a summary of what is known so far from PPH prevention trials.

***Question 1:*** Now that we have reviewed this additional data, do we need to revise the top-ranked threshold from the previous rounds (at least 1000 mL OR signs of haemodynamic instability, whichever comes first)?

***Question 1.1***: If the answer is not to revise, then is it acceptable that the definition (500 mL) and threshold (1000 mL) for triggering action differ?

**<u>Treatments</u>**

**Background**: The experts agreed to an aetiology-based treatment approach, but also agreed that the uterotonic of choice should be administered for all women, regardless of presence of atony.

| Treatment Options | Median |
|---|---|
| Examine and rapidly initiate cause-specific first response (e.g., if trauma: rapid surgical haemostasis; if atony/placental cause: uterotonics and uterine massage) | 9 |
| Tranexamic acid for all women with PPH during CS regardless of aetiology | 8 |
| Plasma expansion with crystalloids for all women with PPH during CS regardless of aetiology | 7.5 |
| Uterotonics for all women with PPH during CS regardless of aetiology | 7 |

***Question 2***: How do we make a recommendation that encompasses these seemingly contradictory statements (i.e., should the first-response uterotonic be administered to all women or just to women with atony?)?

**Background**: The experts agreed that a first response to intraoperative PPH would be to give oxytocin and TXA. However, many women having a caesarean birth would have already received/or be receiving IV oxytocin, and perhaps TXA, for PPH prevention.

| Treatment Options | Median |
|---|---|
| Examine and rapidly initiate cause-specific first response (e.g., if trauma: rapid surgical haemostasis; if atony/placental cause: uterotonics and uterine massage) | 9 |
| Tranexamic acid for all women with PPH during CS regardless of aetiology | 8 |
| Plasma expansion with crystalloids for all women with PPH during CS regardless of aetiology | 7.5 |
| Uterotonics for all women with PPH during CS regardless of aetiology | 7 |

***Question 3***: How do we adapt first-response recommendations if women are already receiving oxytocin infusion (common practice and consensus statement) for PPH prevention?

***Question 4***: In the event that women will routinely be receiving TXA for PPH prevention during CS, what recommendations should we give should they begin to haemorrhage intraoperatively at less than 30 min from the first dose?

**<u>Regarding IV fluids</u>**

**Background**: In the survey, the experts agreed that plasma expansion with crystalloids should be used for all women with PPH during CS, regardless of aetiology.

| Treatment Options | Median |
|---|---|
| Examine and rapidly initiate cause-specific first response (e.g., if trauma: rapid surgical haemostasis; if atony/placental cause: uterotonics and uterine massage) | 9 |
| Tranexamic acid for all women with PPH during CS regardless of aetiology | 8 |
| Plasma expansion with crystalloids for all women with PPH during CS regardless of aetiology | 7.5 |
| Uterotonics for all women with PPH during CS regardless of aetiology | 7 |

***Question 5***: Although we have used "Plasma expansion with crystalloids" in previous surveys, it may not be the best term. Should we use a different term (e.g., increase IV fluids with crystalloids for hemodynamic maintenance)?

***Question 6:*** What should our recommendations be about how to continue IV fluids infusion with crystalloids for hemodynamic maintenance?

***Question 7:*** Do you think that we should be giving more details on type of crystalloids?

**Additional considerations**

***Question 8:*** Are there/should there be additional considerations for any other first-response treatments during the intraoperative period?

**Draft proposed list of postoperative interventions**

**Early detection of PPH and thresholds for triggering first-response management**

- Haemodynamic instability (blood pressure, heart rate, oxygen saturation) with any blood loss volume, OR
- Accumulated blood loss at least 1000 mL (weighted pads + blood loss during CS)

*# If blood loss during CS <500 ml: usual postoperative clinical monitoring.*

*# If blood loss during CS 500-999 ml: further assessment, preparedness, and close monitoring*

**First response treatment**

- TXA (1g in 10 mL IV over 10 min) for all women
- Examine and rapidly initiate cause-specific first response:
  - If trauma suspected: re-laparotomy for surgical haemostasis
  - If atony: Uterotonics
- Uterotonics for all women
- IV fluids with crystalloids for all women

**Pending issues**

**Background**: During the survey, we asked questions about the detection methods, thresholds, and treatments using 2 hours as the postoperative period.

***Question 9***: Is it valid to extend the results to the first 24 hours, or is a different strategy needed for hours 1-2 hours postoperative versus 3-24 hours postoperative?

*For example: Should a woman with cumulative blood loss of 1100 mL in the first 2 hrs have her bleeding managed in the same way as a woman with a cumulative blood loss of 1100 mL in the first 24 hrs?*

**Detection & thresholds**

**Background:** The panel agreed that using **clinical signs of haemodynamic instability** is a more appropriate option for detecting postoperative PPH due to the risk of internal bleeding postoperative.

Additionally, the panel agreed that deploying volumetric methods (e.g., drapes) would be less feasible than monitoring vital signs. Reasons given were a) the possible low acceptability by women (due to discomfort) and b) the lack of feasibility of continued measurement during women's transfer and normal movements.

| | How would you rate each of the methods below for early detection of PPH considering… | | | |
|---|---|---|---|---|
| **Blood loss measurement and other PPH detection methods** | **usefulness?** | **feasibility?** | **acceptability?** | **resources required?** |
| Clinical signs of haemodynamic instability | 8.0 | 8.0 | 8.0 | 3.5 |
| Volumetric/gravimetric + clinical signs of haemodynamic instability | 7.0 | 7.0 | NA | NA |
| Clinical judgement such as rate of flow and duration | 5.0 | 6.0 | 7.0 | 2.0 |
| Gravimetric | 5.0 | 3.0 | 5.0 | 7.0 |
| Volumetric | 5.0 | 4.5 | 7.0 | 7.0 |
| Visual estimation of blood loss | 4.0 | 7.0 | 7.0 | 1.0 |
| Visual charts and early warning scores (EWS) | 7.0 | 7.0 | 6.5 | 6.0 |
| Visual estimation + visual charts /EWS | 7.0 | 7.0 | NA | NA |
| Volumetric + gravimetric | 7.0 | 5.0 | NA | NA |

Given the diverse realities of postoperative monitoring, priority should be given either to frequent monitoring of vital signs or the use of continuous monitoring devices, dependent on the availability of equipment and personnel.

***Question 10***: Consequently, what kind of guidance should we give, if any? How frequently should the woman's haemodynamic status be monitored? For how long? With what kind of devices?

**Background**: In addition to monitoring vital signs, providers should be **monitoring blood loss.** The experts agreed that in the postoperative period, providers should continue with the cumulative measurement of blood loss (i.e., adding intraoperative blood loss to postoperative blood loss to calculate total loss).

| | Median |
|---|---|
| The volume of intraoperative blood loss should be taken into consideration when determining whether the postoperative threshold had been reached | 8 |

***Question 11:*** What guidance should we give to reflect cumulative intra- and postoperative blood loss?

We propose, **as an example**, the following approach to blood loss monitoring **in situations in which the woman's vital signs are stable**. ***If vital signs indicate haemodynamic instability, the guidance should be to initiate treatment regardless of postoperative blood loss.***

| Intraoperative | | Postoperative | | |
|---|---|---|---|---|
| **Blood Loss** | **Action Indicated** | **Type of Monitoring** | **Cumulative Blood Loss** | **Action Indicated** |
| <500 | No treatment | Normal monitoring | Cumulative blood loss <1000 | Do not initiate treatment. Continue normal monitoring |
| 500-999 | No treatment | Further assessment, preparedness, and more frequent postop monitoring | Cumulative blood loss <1000 | Do not initiate treatment. Return to normal monitoring |
| 500-999 | No treatment | Further assessment, preparedness, and more frequent postop monitoring | Cumulative blood loss >1000 | Initiate treatment, and monitor the woman's response to the treatment |
| ≥1000 | Initiate treatment and monitor woman's response | Monitor woman's response to treatments | - | - |

**DAY 2: 28 September 2022**

***Question 12:*** How should we frame the first-response strategies? Or bundle?

- Algorithm?
- Checklist?
- Or a hybrid?

***Question 13:*** Should we include preparedness interventions at the facility level?
**Prepare for CS PPH**

Blood loss measurement

- Calibrated containers
- Pump/aspirator/vacuum suction
- Pads/swabs/lap cloths, etc. with known dry weight
- Scale to weigh the above

Medications

- TXA
- Uterotonics (Oxytocin, ergometrine, sulprostone)
- Crystalloids

Haemodynamic Monitoring

- CRADLE device/ cardiac monitor/ oximeter/sphygmomanometer?

**Supplementary File S4.** List of contributors

**EXPERTS**
Fadhlun ALWY AL-BEITY - Tanzania
Edgardo ABALOS - Argentina
Sabaratnam ARULKUMARAN - UK
Nabhan ASHRAF - Egypt
Brendan CARVALHO - USA
Catherine DENEUX-THARAUX - France
Alexander DUMONT – France
Maria Fernanda ESCOBAR VIDARTE - Colombia
Cherrie EVANS - USA
Sue FAWCUS - South Africa
Hadiza GALADANCI - Nigeria
Justus HOFMEYR - South Africa
Caroline HOMER - Australia
Tippawan LIABSUETRAKUL - Thailand
Pisake Lumbiganon - Thailand
Elliott MAIN - USA
Judith Maua ONG'AYI - Kenya
PHAN NGUYEN Quoc Thuan - Vietnam
Zahida QURESHI - Kenya
John VARALLO - USA

**OBSERVERS**
Arri COOMARASAMY - UK
Inês NUNES - Portugal
Andrew WEEKS - UK
Claudio SOSA - Uruguay

**STEERING GROUP**
Fernando ALTHABE - Argentina
Suellen MILLER - USA
Veronica PINGRAY - Argentina
Caitlin R. WILLIAMS - USA

**WHO SECRETARIAT**
Ioannis GALLOS
Olufemi OLADAPO
Mariana WIDMER
